# Estimating the Effects of Nordic Diets on the Risk of Major Adverse Liver Outcomes: a Target Trial Emulation across Two Cohorts in Sweden

**DOI:** 10.64898/2026.01.07.26343586

**Authors:** Michael Fridén, Conor J. MacDonald, Hannes Hagström, Agneta Åkesson, Søren Nielsen, Daniel B. Ibsen

**Author notes:** **Corresponding author:** Michael Fridén.

## Abstract

**Background:** Randomized trials have shown that a healthy Nordic diet (HND) improves liver steatosis, but there is limited evidence on the effects of Nordic dietary patterns on the risk of major adverse liver outcomes (MALO). We specified a hypothetical target trial protocol to estimate the effects of adhering to a HND or the Nordic Nutrition Recommendations 2023 (NNR23) on the 24-year risk of MALO in a middle-aged to elderly Swedish population.

**Methods:** Two pooled population-based cohorts including n=64,406 men and women (Cohort of Swedish Men (COSM) and the Swedish Mammography Cohort (SMC)) with repeated measurements on diet and confounders in 1997, 2008/2009 and 2019 were used to emulate population-adapted versions of the diets. Under the assumptions of no unmeasured confounding, selection bias or measurement error, the parametric g-formula was used to estimate 24-year risks of MALO from each hypothetical intervention. Secondary analyses included comparing the HND and NNR23 with a low-adherence group; reducing alcohol as an additional hypothetical intervention; and assessing risk of all-cause mortality.

**Results:** The estimated 24-year risk of MALO in the HND was 0.53% (95% CI: 0.38, 0.73), in the NNR23 diet 0.70% (95% CI: 0.57, 0.90) and in no intervention 0.64% (95% CI: 0.56, 0.77). Estimated risk differences (RDs) of MALO for HND versus no intervention and NNR23 versus no intervention were -0.11% (95% CI: -0.27, 0.07) and 0.06% (95% CI: -0.04, 0.15), respectively. Compared to NNR23, the estimated RD for the HND was -0.17% (95% CI: -0.38, 0.05). Meaningful risk reductions following the HND were estimated when compared to a low-adherence diet group (-1.50% (95% CI: -9.53, -0.05)), when including reducing alcohol, and for all-cause mortality (-2.67% (95% CI: -3.51, -1.85) versus no intervention; -1.68% (95% CI: - 2.75, -0.62) versus NNR23)).

**Conclusion:** We estimated no clear risk reductions from a population-adapted HND or a NNR23 diet on the 24-year risk of MALO when compared to each other or no intervention. However, when either compared to a low-adherence group or when including reducing alcohol as a hypothetical intervention or when specifying all-cause mortality as the outcome, we estimated meaningful risk differences following the HND.

## Background

The healthy Nordic diet (HND) emphasizes foods traditionally consumed by the Nordic populations and closely aligns with the Nordic Nutrition Recommendations 2023 (NNR23) [1]. A clear distinction, however, is the focus on specific Nordic foods in the HND, where e.g., rapeseed oil is emphasized over olive oil, oats and rye over wheat and rice and pears and apples over oranges and bananas [1–3]. High adherence to the HND has been associated with a lower risk of various cardiometabolic diseases, including type 2 diabetes (T2D), cardiovascular disease, cancer as well as all-cause mortality [4,5].

In a recent 12-month randomized controlled trial (RCT), a HND was shown to reduce liver fat and improve liver function markers in Swedish adults with T2D or prediabetes, when compared to a group assigned to the previous version of the NNR [6]. Yet, it remains unknown whether short-term effects on intermediate risk markers translate to lower risk of more advanced stages of liver diseases, such as cirrhosis, complications of cirrhosis and hepatocellular carcinoma (HCC). This is important as the incidence of metabolic dysfunction-associated steatotic liver disease (MASLD), alcohol-related cirrhosis, unspecified cirrhosis (likely reflecting MASLD-related cirrhosis), and HCC, all potentially modifiable by lifestyle factors, has increased in Sweden since the early 2000s [7]. Conversely, compensated cirrhosis, i.e., the stage of cirrhosis where the liver still functions, is often asymptomatic, making most patients unaware of its presence. Once the liver decompensates, long-term survival prognosis decreases markedly [8], emphasizing the importance of effective population-level preventive strategies. Targeting diet, through Nordic diets, could potentially be one promising strategy. However, obtaining such level of evidence would ideally require an RCT with over 20 years of follow-up, given that cirrhosis often takes decades to develop [9]. A trial like that would likely be unfeasible to conduct, which is why observational studies are necessary.

Due to a number of methodological issues, however, current observational analyses examining dietary patterns and incidence of more advanced stages of liver diseases do not provide answers comparable to those that could be obtained from a RCT. Conventional nutritional epidemiological studies often compare those with the highest adherence to a dietary pattern to those with the lowest, typically based on a single time point, not truly reflecting how a trial would have been designed, if feasible. In contrast, a well-designed RCT would clearly specify the intervention and comparator arm, inclusion and exclusion criteria, the causal contrast, and thoroughly define how protocol deviations would be handled. Newer causal inference frameworks, such as the target trial emulation framework, have been developed to enable researchers to ask more meaningful causal questions and analyze them in a way that mimics the conditions of a RCT [10–13].

As such, we used the target trial framework to emulate a hypothetical target trial comparing sustained adherence to the HND as well as a diet aligning with the NNR23 to each other and to no intervention, in relation to the 24-year risk of major adverse liver outcomes (MALO) in middle-aged to elderly Swedish adults.

## Methods

### Step 1: Specification of the target trial

To estimate the effects of sustained adherence to a HND or the NNR23 versus each other or no intervention (i.e., the observed diet over time) on the 24-year risk of MALO in Swedish adults, a three-arm parallel-designed *ad libitum* pragmatic randomized trial would be conducted. A hypothetical target trial protocol is shown in **Table 1**.

**Table 1.** Target trial emulation protocol of two Nordic diets on the risk of MALO.

| Protocol | Target trial | Target trial emulation |
| --- | --- | --- |
| **Eligibility criteria** | Recruited in 1997, aged $\geq 45$ years, no previous chronic liver diseases (CLDs), including MALO, and no history of cancer. | Same as the target trial + valid information on baseline diet and covariates available. |
| **Treatment strategies** | Participants are randomized into three groups:<br>1) Participants receive information about the HND as well as tailored dietary advice as how to reach the HND guidelines<br>2) Participants receive information about the NNR 2023 guidelines as well as tailored dietary advice as how to reach the NNR 2023 guidelines<br>3) No intervention<br>Intervention thresholds for the HND are:<br>• at least 2 servings of whole-grain bread (including crisp bread)/day<br>• at least 3 servings of oatmeal or rye porridge/week<br>• at least 250 grams of Nordic fruits, vegetables and berries per day (e.g., cabbage, peas, pears, apples, blueberries, raspberries)<br>• at least 3 servings of fatty fish (e.g., salmon, mackerel, herring)/week<br>• $\leq 280$ grams of red meat/week<br>• $\leq 70$ grams of processed red meat/week<br>• using rapeseed oil in both cooking and in salad dressing<br>Intervention thresholds for the NNR23 diet are:<br>• $\leq 4$ cups of coffee/day<br>• at least 90 grams of whole-grain cereals/day<br>• at least 500 grams of fruits, vegetables and berries/day<br>• at least 100 grams of pulses/day<br>• at least 20 grams of nuts/day | Same as the target trial + the assumption that reported intake from FFQs represent the average intake until answering the next FFQ, loss to follow-up or end of follow-up. The no intervention arm will assume that individuals eat according to their reported intake from the FFQ.<br>Population-adapted intervention thresholds for the HND are:<br>• at least 2 servings of whole-grain bread (including crisp bread)/day<br>• at least 0.5 servings of oatmeal or rye porridge/week<br>• at least 125 grams of Nordic fruits, vegetables and berries per day (e.g., cabbage, peas, pears, apples, blueberries, raspberries)<br>• at least 0.5 servings of fatty fish (e.g., salmon, mackerel, herring)/week<br>• $\leq 420$ grams of red meat/week<br>• $\leq 350$ grams of processed red meat/week<br>• using either rapeseed oil in cooking or in salad dressings<br>Population-adapted intervention thresholds for the NNR23 diet are:<br>• $\leq 4$ cups of coffee/day<br>• at least 90 grams of whole-grain cereals/day<br>• at least 250 grams of fruits, vegetables and berries/day<br>• at least 10 grams of pulses/day<br>• at least 0.5 servings of fatty fish/week<br>• at least 0.5 servings of other fish/week<br>• $\leq 420$ grams of red meat/week |
|  | • at least 2 servings of fatty fish/week<br>• at least 1 serving of other fish/week<br>• at least 25 grams of vegetable oil/day<br>• $\leq 280$ grams of red meat/week<br>• $\leq 70$ grams of processed red meat/week<br>• $\geq 350$ grams of low-fat dairy/day<br>• $\leq 130$ grams of potatoes/day<br>• no more than 1 egg/day<br>• no more than 350 grams of poultry/week<br>• no more than 100 grams of juice/day<br>Individuals would be encouraged to continue to eat according to the intervention until the next scheduled visit. If an individual develops an intolerance or allergy to any of the foods they will be exempt from making that change. | • $\leq 350$ grams of processed red meat/week<br>• $\geq 100$ grams of low-fat dairy/day<br>• $\leq 130$ grams of potatoes/day<br>• no more than 1 egg/day<br>• no more than 350 grams of poultry/week<br>• no more than 100 grams of juice/day<br>If the FFQ indicates that an individual is below or above the recommended value, the value is changed to a value compatible with the intervention.<br>Participants who develop intolerances or allergies to included foods are not exempt from making changes to that food in the main analysis (only in sensitivity analysis). |
| **Assignment procedure** | Random assignment to the HND, the NNR 2023 diet or no intervention. Participants would be aware of their allocated diet. | We assume randomization conditional on the following baseline covariates: age, sex, education, smoking, alcohol intake, physical activity, sleep, dietary supplements use, BMI, history of hypertension, hypercholesterolemia, diabetes, IBS or IBD. |
| **Outcomes** | Primary outcome: 24-year risk of MALO defined as a composite outcome of decompensated cirrhosis (portal hypertension, ascites (in conjunction with any CLD), hepatorenal syndrome or bleeding esophageal varices), liver transplantation, or HCC (including death from these outcomes).<br>Secondary outcome: 24-year risk of all-cause mortality. | Same as the target trial. |
| **Follow-up** | From randomization in 1997 to outcome, non-MALO death, loss to follow-up or administrative end of follow-up (24 years after baseline), whichever occurs first. | Same as the target trial + baseline is defined as January 1 1998 when all participants had returned their food-frequency questionnaire (FFQ). Loss to |
|  |  | follow-up is defined as not returning the FFQ in 2009 or 2019. |
| **Causal contrast** | Intention-to-treat effect and the per-protocol effect (total effect). | Observational analogue of the per-protocol effect (total effect). |
| **Statistical methods** | Intention-to-treat: parametric g-formula with an indicator for intervention group, adjustment for variables associated with loss to follow-up.<br>Per-protocol effect: parametric g-formula with an indicator for intervention group, adjustment for pre- and post-baseline variables associated with adherence to the protocol and/or loss to follow-up. | Same as the target trial for the per-protocol effect. |

## Eligibility criteria

Eligible to participate in the trial would be men and women over the age of 45 with no history of cancer, MASLD or other chronic liver diseases (CLDs), including MALO, when recruited in 1997.

## Outcome and follow-up

The primary outcome would be MALO, a composite outcome consisting of decompensated cirrhosis (hepatic ascites, portal hypertension, hepatorenal syndrome and bleeding esophageal varices), hepatocellular carcinoma (HCC) and liver transplantation, whichever comes first. Ascites requires a concomitant diagnosis of CLD (e.g., alcoholic-liver disease cirrhosis or chronic viral hepatitis) (**Supplementary Table 3**). This requirement exists because ascites can result from other non-hepatological conditions, such as heart failure and non-HCC malignancies. Participants would be followed from randomization (1998-01-01) to incident MALO, loss to follow-up, death or administrative end of follow-up (2021-12-31), whichever comes first. A secondary outcome would be all-cause mortality.

## Treatment strategies

Diet strategies would be randomly allocated in a 1:1:1 ratio. Participants would be aware of their allocated diet and they would be followed up each month the first year and each 6 months thereafter to ensure good protocol adherence. Participants allocated to the HND would be given advice to eat the following: ≥2 servings of whole-grain bread/day; ≥3 servings of oatmeal or rye porridge/week; ≥250 grams of Nordic F&V/day, such as apples, pears, berries and cabbage; ≥3 portions of fatty fish/week; and ≤350 grams of red meat/week, of which ≤70 grams would come from processed red meat. As the HND heavily emphasizes rapeseed oil as the main fat source, participants would be advised to use only rapeseed oil in cooking and as base in salad dressings. The foods included in the HND are inspired by the Healthy Nordic Food Index (HNFI) developed by Kyrø et al., 2013 [14], with the addition of rapeseed oil and red- and processed red meat as the HND is primarily plant-based. Food thresholds specified for the HND are also commonly used in randomized trials, and align well with the NNR 2023 [1–3,15]. At each monthly meeting, if a participant would report consuming less than the lower threshold value of a food to increase, the participant would be advised to consume an amount aligned with the lower bound of that threshold, e.g., if reporting a value of 200 grams of Nordic F&V/day, advice would be given to consume exactly 250 grams/day. If a participant would report an amount above the threshold value, no intervention would be given for that particular food, but rather advised to continue consuming the same amount. The reverse would apply for foods to decrease. Such intervention strategies are called threshold interventions [16].

Participants allocated to the NNR23 diet would be given advice to consume the following: ≤ 4 cups of coffee/day; ≥90 grams of whole-grain cereals/day; ≥500 grams of F&V/day; ≥100 grams of pulses/day; ≥3 portions of fish per week, of which ≥2 portions would be fatty fish; ≥350 grams of low-fat dairy/day; ≤130 grams of potatoes/day; ≥25 grams of vegetable oil/day; ≥20 grams of nuts/day; ≤350 grams of red meat/week, of which ≤70 grams would come from processed red meat; ≤1 egg/day; ≤350 grams of poultry/week; and ≤100 grams of juice/day. Thresholds are based on the NNR 2023 combined with the development of the NNR23 dietary pattern score [15,17].

No advice would be given to participants allocated to the no intervention group.

Due to ethical reasons, if a participant would develop an allergy or intolerance to milk, gluten or nuts before each meeting, they would be exempt from making changes to that particular food.

## Causal contrast and statistical analysis

The primary causal contrast of interest in the target trial would be the per-protocol effect, i.e., the joint effect of adhering to the assigned protocol over the full intervention period. Due to the longer follow-up of 24 years, a large proportion of participants would probably deviate from the protocol as time passes, making the intention-to-treat effect (i.e., the effect of treatment assignment) less valuable for public health decision making [18]. The intention-to-treat effect would be specified as a secondary causal contrast.

The per-protocol effect may be estimated using the parametric g-formula, under assumptions of no unmeasured confounding, no selection bias, no measurement errors and no model misspecifications [19]. The parametric g-formula is a subclass of g-methods (a family of statistical methods for causal inference) used to obtain unbiased effect estimates of time-varying treatments in the presence of treatment-confounder feedback. Since only the initial treatment assignment would be randomized at baseline, and not adherence to the protocol thereafter, time-varying covariates influenced by diet, such as body mass index (BMI) or hyperlipidemia, are likely to affect adherence at subsequent time points, thereby creating treatment-confounder feedback. Conventional methods such as outcome regression adjustment would not suffice to account for this feedback, but g-methods would, as previously described in detail [19]. In short, the parametric g-formula is a generalization of standardization that allows for adjustment of time-varying confounding affected by prior exposure by modeling, parametrically, the joint distributions of outcomes, competing outcomes, exposures and covariates under hypothetical interventions (e.g., had everybody adhered to all dietary components in the HND protocol versus no intervention). Monte Carlo simulation is then used to repeatedly sample from the fitted parametric models across time points to generate predicted outcomes under each hypothetical intervention. Risk differences (RDs) and risk ratios (RRs) are then calculated from standardized risks of the outcome and percentile-based 95% CIs are retrieved from bootstrapping with n=500 resamples.

As a secondary analysis, causal survival forests would be used to estimate conditional average treatment effects (CATEs) over a set of prespecified predictor variables [20]. In short, a causal forest builds trees based on splits of the covariates that maximize treatment effect heterogeneity, leaving both treated and controls in separate, so-called, leaves. These trees, and thus leaves, are then aggregated into a forest and averaged to estimate how treatment effects vary across individuals.

### Step 2: Emulation of the hypothetical target trial

To emulate the hypothetical target trial, we used data from two population-based cohort studies in Sweden; the Cohort of Swedish Men (COSM) and the Swedish Mammography Cohort (SMC). Both cohorts belong to the National Research Infrastructure: Swedish infrastructure for medical population-based life-course and environmental research (SIMPLER) (http://www.simpler4health.se/). SMC and COSM were conducted in accordance with the declaration of Helsinki and were approved by the Regional Ethical Review Board at Karolinska Institutet, Stockholm, Sweden.

The TARGET (TrAnsparent ReportinG of observational studies Emulating a Target trial) guidelines for transparent reporting were adhered to, and the corresponding checklist is available as **Supplementary Material** [13].

## Observational data

### COSM and SMC

SMC and COSM were initiated in 1987 and 1997, respectively, with the aim to investigate associations between dietary exposures and chronic disease outcomes [21]. Participants in both cohorts are followed up every decade with both lifestyle questionnaires and food frequency questionnaires (FFQs). Information on diet and lifestyle habits from 1997, 2008/2009 and 2019 were used for this study. A total of n=156 333 individuals from both cohorts were asked to participate in 1997, of which n=88 077 agreed. After excluding individuals at baseline with incorrect or missing personal identification number, other missing data, missing data on baseline covariates, energy misreporting (defined as within 3 standard deviations of total mean energy intake on the log-scale), diagnosis of any CLDs (including MALO) or cancer, n=64 406 participants remained. A complete flow-chart is shown in **Supplementary Figure 1**.

Baseline diet (including alcoholic beverages) in 1997 was assessed using a validated FFQ, containing 96 items with response categories ranging from never to ≥ 3 times/day. The FFQ also contained open-ended questions on foods and beverages typically consumed in Sweden, such as dairy, bread, coffee and beer. Average intakes of foods in grams were calculated by multiplying the frequency of each food item with age- and sex-specific portion sizes estimated from weighed food records in a subgroup of the cohorts. Correlation coefficients between the FFQ and 7-day food records were 0.7 for fermented milk, 0.3-0.7 for red and processed red meat, 0.5-0.7 for fruits, 0.4-0.6 for vegetables and 0.5-0.7 for whole grains [22,23]. Follow-up FFQs in 2009 and 2019 were extended to contain 132 items, capturing new food items on the market. Missing data on individual foods were assumed to be zero consumption. Categorizations of foods are presented in **Supplementary Table 2**.

Potential time-fixed and time-varying confounders were determined *a priori* based on the background literature, subject-matter expertise and with the use of a directed acyclic graph (DAG) (see **Supplementary Figure 2** for a simplified version) [24]. Time-fixed confounders were age, sex and education. Time-varying confounders were BMI, history of hyperlipidemia, hypertension, diabetes, irritable bowel syndrome (IBS) or inflammatory bowel disease (IBD), sleep, smoking, physical activity, alcohol intake, use of dietary supplements and intake of Nordic foods. IBS and IBD (yes/no) were diagnosed through the Swedish National Patient Register (ICD-9: 555, 556, 564.1 and ICD-10: K58, K50, K51). All other confounders were assessed using self-reported lifestyle questionnaires. Education was categorized into <9 years, 9-12 years or >12 years (University) in school; smoking was categorized into never, former or current smoker and physical activity was categorized into <1 hour/week, 1 hour/week, 2-3 hours/week or >3 hours/week of exercise. History of diabetes, hyperlipidemia and hypertension were coded as yes/no. Sleep (hours/day), alcohol (g/day) and Nordic foods (g/day) were measured as continuous variables. BMI was calculated by dividing self-reported weight in kg by self-reported height in cm squared.

Incident cases of MALO were ascertained using the Swedish National Patient Register (from inpatient stays and from outpatient care since 2001), the Swedish Cancer Register and the Swedish Cause of Death Register. MALO was defined in accordance with *International Classification of Diseases, Tenth Revision* (ICD-10) (**Supplementary Table 3**). Both main and contributing diagnoses were included, meaning that we used all diagnostic codes recorded during a hospital contact. Positive predictive values of MALO in Swedish registers range from 84% for HCC to 96% for esophageal varices [25]. Ascites combined with a code for CLD has a positive predictive value of 91% [25]. All-cause mortality was ascertained from the Swedish Cause of Death Register.

## Modifications to the target trial protocol

In addition to the specified eligibility criteria in the target trial, participants had to have valid information on diet (defined as within 3 standard deviations of total mean energy intake on the log-scale) and complete data on baseline covariates (i.e., diet and all confounders listed below). Randomization was emulated by adjusting for the following baseline confounders: age, sex, education, smoking, alcohol intake, physical activity, sleep, use of dietary supplements, BMI, history of hypertension, hypercholesterolemia, diabetes, IBS or IBD. A simplified DAG depicting our causal assumptions can be found in **Supplementary Figure 2**. We assumed that reported intakes from each FFQ represent the average intakes until answering the next FFQ. The no intervention arm assumes that individuals eat according to their reported intakes from the FFQs.

As no one adhered to all components of the HND or the NNR23 guidelines over follow-up, thereby risking violating the positivity assumption (i.e., the non-zero probability of receiving either intervention), population-adapted thresholds had to be specified (**Supplementary Table 1**). Lower thresholds had to be specified for F&V, pulses, fatty fish, other fish, red meat, processed red meat, low-fat dairy, oatmeal or rye porridge, Nordic F&V and rapeseed oil.

Furthermore, as few individuals consumed >0 grams of nuts and vegetable oil, these foods were excluded from the NNR23 diet (**Supplementary Table 1**). Due to the strong underlying assumptions described under causal contrasts and statistical analysis, participants were not exempt from making changes to their diet if developing an intolerance or allergy to the food, in the primary analysis (although included in sensitivity analyses). Contrasts in all-cause mortality risks were only calculated for the comparison between the HND and no intervention and the HND and NNR23. Lastly, participants were followed up every 10^th^ year (2009 and 2019) instead of each month to every 6 months. Follow-up started 1^st^ of January 1998 when all had returned their FFQ. Loss to follow-up was defined as not returning the FFQ in 2009 or 2019.

## Causal contrast and statistical analysis

Estimating the observational analogue of the per-protocol effect on the 24-year risk of MALO from contrasting a HND and the NNR23 diet with each other and with no intervention requires adjusting for the following baseline covariates: age, sex and education and the following time-varying covariates: BMI, history of hyperlipidemia, hypertension, diabetes, IBS or IBD, sleep, smoking, physical activity, alcohol intake, use of dietary supplements and intake of Nordic foods. To estimate total effects of the diets, competing events (deaths from other causes) were not treated as censoring events [26]. Missing values in covariates over the two follow-up time points (ranging from 1.9-2.3% for physical activity to 8.9-11.9% for the use of supplements) were replaced with the last observed value. The SMC cohort and the COSM cohort were pooled due to few events in each cohort. Risks were estimated using the parametric g-formula under the assumption that the measured covariates are sufficient to adjust for both confounding and selection bias due to loss-to follow-up (i.e., that sequential exchangeability holds at each time point) and then compared on both the difference and ratio scale. Details on the functional forms of the covariates are provided in **Supplementary Table 4**. R (version 4.3.1) was used to compute all statistical analyses and the following packages were used: “*gfoRmula*”, “*grf*”, “*mice*”, “*Hmisc*”, “*tidyverse*” and “*here*”.

Secondary analyses included:

- Estimating the effects of the HND on all-cause mortality as a secondary outcome.
- Contrasting the HND and the NNR23 diet with a low-adherence diet group. The low-adherence diet was defined as high intakes of red and processed red meat, and low intakes of F&V and whole-grain cereals. Threshold interventions for these foods were defined as reversing current dietary recommendations laid out in NNR 2023, such that ≥ 500 grams of F&V/day became ≤500 grams of F&V/day. For red meat, the threshold intervention was ≥280 grams/week, for processed red meat ≥70 grams/week and for whole-grain cereals ≤90 grams/day.
- Contrasting the HND and the NNR23 diet with each other and with no intervention, but now including reducing alcohol as an additional hypothetical threshold intervention (< 0.5 units (or 6 grams)/day). Alcohol is a potential driving cause of MALO and as the NNR 2023 emphasize reducing alcohol to zero, this secondary analysis was deemed reasonable to conduct, although not a part of the food-based dietary guideline.
- Intervening on all but one food component at a time for both the NNR23 diet and the HND. To reduce computational burden, six foods per diet previously associated with liver disease were excluded from being intervened on [27–34].
- Exempting participants with food intolerances or allergies from being intervened on that particular food. Participants had to answer whether they actively exclude dairy or gluten from their diet at each follow-up. We made the strong assumption that the reason for the exclusion was due to intolerance or allergy. No information on food restrictions was provided at baseline, hence a further strong assumption was that no participants excluded any foods at baseline. Due to the above, we did not specify this analysis as the primary analysis.
- Changing the thresholds for low-fat dairy (from ≥100 to ≤500 grams per day), potatoes (from ≤130 to ≥50 grams per day) and whole-grain cereals (from ≥90 to ≥110 grams per day) in the NNR23 diet. The reason being that both low-fat dairy and potatoes are specified in ranges (50-130 grams for potatoes in the development of the NNR23 diet score and 350-500 grams for low-fat dairy in NNR 2023) and that some whole-grain cereal products may not be 100% whole-grains.
- Conducting conventional time-updated analyses using pooled logistic regression with high vs low adherence of a HND or NNR23 as the exposure.
- Using causal survival forests to examine effect heterogeneity of comparing high versus low adherence groups of the HND at baseline on the 12-year mean survival time of MALO and all-cause mortality [20]. Potential effect modifiers were all baseline confounders included in the main analysis. Average treatment effects (ATEs) were estimated across tertiles of predicted CATEs, hereafter termed effect scores, using the doubly-robust Augmented Inverse Propensity Weighting (AIPW) estimator. A detailed description of the causal forest methodology is found in **Supplementary Material**.

Several sensitivity analyses were prespecified including:

- Adjusting for total energy intake. This adjustment would change the research question to an isocaloric setting instead of an *ad libitum* setting, but arguments have been put forth that not adjusting for total energy- or total food intake may introduce confounding bias from unknown or unmeasured determinants of dietary intake [35,36].
- Varying the functional forms of included covariates to examine robustness to model misspecification.
- Varying the order of covariates in the models to examine robustness to model misspecification.
- Using external causes of mortality as negative control outcomes (ICD codes: V01–Y98, except intentional self-harm: X60–X84) to examine robustness to unobserved/residual confounding. The assumption being that MALO and external causes of mortality share the same confounding structure but no true causal effects from diets on the negative control outcome are expected [37].
- Missing data at follow-up were handled through multivariate imputation of chained equations (MICE) with n=1 imputations [38].

## Results

Out of n=156 333 individuals invited, n=88 077 agreed to participate. After initial exclusion of participants with incorrect or missing ID number, other missing data or history of cancer (in the COSM cohort), n=84 890 remained with complete data on diet in 1997. Out of these, n=20 484 were further excluded due to missing information on covariates, history of cancer, prevalent MASLD and other CLD, energy misreporting and death before 1 January, 1998, leaving a final pooled study sample of n= 64 406 (**Supplementary Figure 1**). Participants had a median age of 59 years (IQR: 53-68), a BMI of 25.0 kg/m^2^ (IQR: 23.0-27.4), 23.2% were current smokers, 18.5% had a university degree and 4.9% had diabetes (**Table 2**). Baseline characteristics stratified by dietary adherence are found in **Supplementary Table 6**. Over the 24-year follow-up, n=405 cases of MALO were reported (n=119 of HCC, n=18 of HCC deaths, n=6 of liver transplantations, n=247 of decompensated cirrhosis and n=15 of deaths thereof). A total of n=29 255 died of any cause. Out of these, n=468 died of external causes.

**Table 2.** Baseline characteristics of the pooled cohort^1^.

|  | Study population (n=64 406) |
| --- | --- |
| Sex (n (%) women) | 29 229 (45.4) |
| Age (years) | 59 (53-68) |
| University degree (n (%)) | 11 902 (18.5) |
| Smoking (n (%) current) | 14 970 (23.2) |
| Physical activity ($\geq 2$ hours/week of exercise) | 37 212 (57.8) |
| Sleep (hours/day) | 7 (7-8) |
| Use of supplements (n (%) yes regularly) | 12 705 (19.7) |
| BMI (kg/m<sup>2</sup>) | 25.0 (23.0-27.4) |
| Hypertension (n (%)) | 13 999 (21.7) |
| Hypercholesterolemia (n (%)) | 7600 (11.8) |
| Diabetes (n (%)) | 3156 (4.9) |
| IBS or IBD (n (%)) | 331 (0.5) |
| Total energy intake (kcal/day) | 2126 (1649-2740) |
| Alcohol intake (g/day) | 5 (1-11) |
| Coffee (g/day) | 552 (354-820) |
| Whole-grain bread (g/day) | 94 (53-154) |
| F&V (g/day) | 290 (192-422) |
| Pulses (g/day) | 19 (16-24) |
| Fatty fish (g/day) | 9 (6-15) |
| Other fish (g/day) | 10 (8-25) |
| Red meat (g/day) | 45 (28-66) |
| Red processed meat (g/day) | 32 (19-45) |
| Low-fat dairy (g/day) | 192 (19-397) |
| Potatoes (g/day) | 99 (65-129) |
| Egg (g/day) | 8 (3-16) |
| Poultry (g/day) | 8 (8-10) |
| Juice (g/day) | 14 (0-45) |
| Oatmeal/rye porridge (g/day) | 10 (0-43) |
| Nordic F&V (g/day) | 123 (74-185) |
| Whole-grain cereals (g/day) | 128 (73-218) |
| Rapeseed oil (n (%) yes in cooking or salad dressing) | 11 975 (18.6) |

<sup>1</sup>Values are presented as median (IQR) for continuous variables and as count (%) for categorical variables.
BMI, Body Mass Index; IBD, Inflammatory Bowel Disease; IBS, Irritable Bowel Syndrome.

## Nordic diets and MALO

The estimated 24-year risk of MALO in the HND was 0.53% (95% CI: 0.38, 0.73), in the NNR23 diet 0.70% (95% CI: 0.57, 0.90) and in no intervention 0.64% (95% CI: 0.56, 0.77). Estimated RDs of MALO for the comparisons between the HND and no intervention and the NNR23 diet and no intervention were -0.11% (95% CI: -0.27, 0.07) and 0.06% (95% CI: -0.04, 0.15), respectively. Corresponding RRs were 0.83 (95% CI: 0.59, 1.11) and 1.09 (95% CI: 0.94, 1.22). Compared to NNR23, the estimated RD for the HND was -0.17% (95% CI: -0.38, 0.05) and the RR was 0.76 (95% CI: 0.53, 1.08) (**Figure 1**). The average proportion of individuals who would have been required to change their diet at each time point to adhere to the diet strategy was 93.89% for the HND and 96.10% for the NNR23 diet (**Supplementary Table 7**).

**Figure 1.**
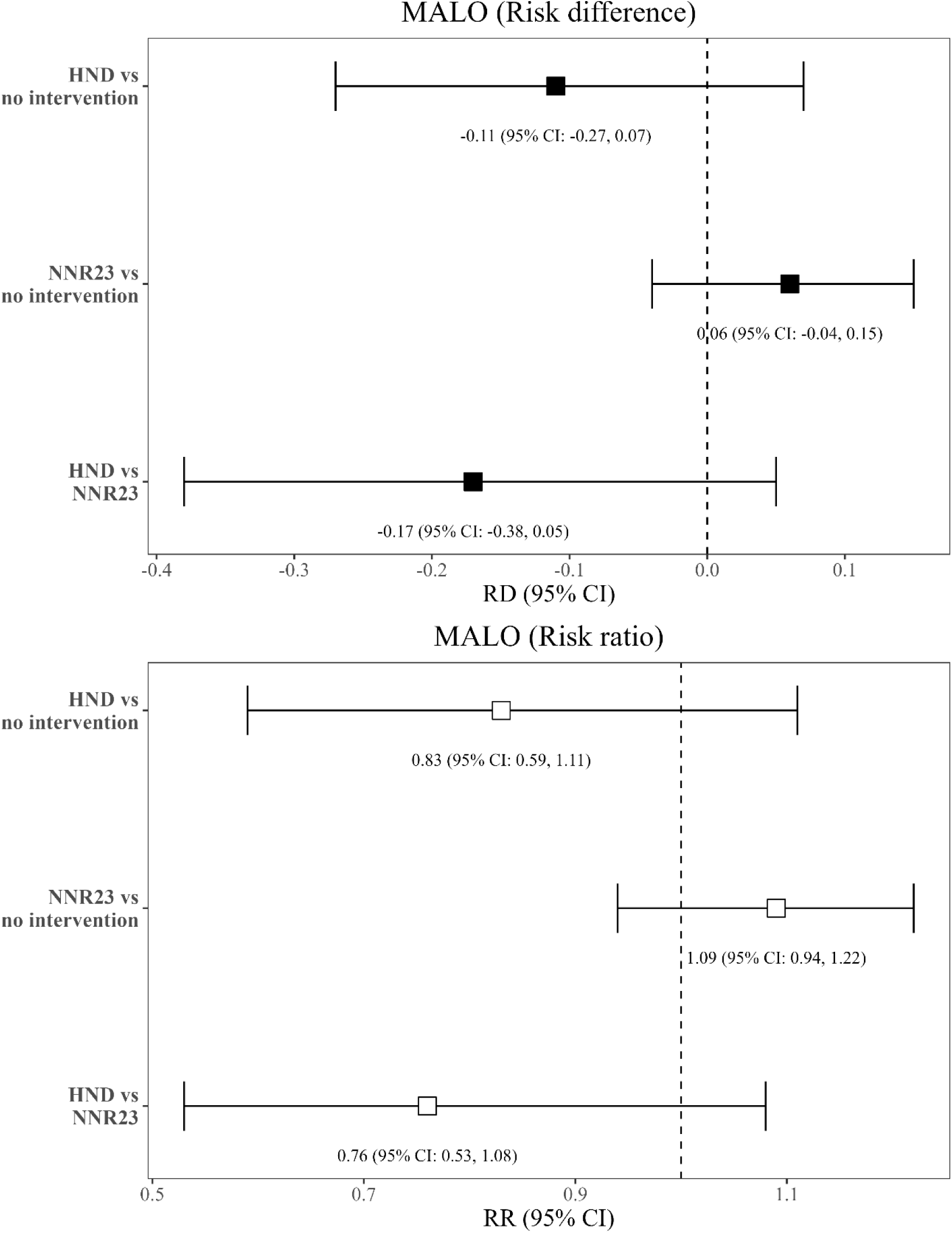
Nordic diets and risk of MALO. Estimates are based on a simulated data set of n=64 406 participants using the parametric g-formula and are adjusted for age, sex and education at baseline and BMI, history of hyperlipidemia, hypertension, diabetes, IBS or IBD, sleep, smoking, physical activity, alcohol intake, use of dietary supplements and intake of Nordic foods as time-varying covariates, and their histories. HND, Healthy Nordic Diet; MALO, Major Adverse Liver Outcomes; NNR23, Nordic Nutrition Recommendations 2023; RD, Risk Difference; RR, Relative

## The HND and all-cause mortality

For all-cause mortality, the estimated 24-year risk was 39.9% (95% CI: 39.1, 40.9) for the HND, 41.6% (95% CI: 40.7, 42.5) for the NNR23 diet and 42.5% (95% CI: 42.0, 43.3) for the no intervention group. The estimated RD for the HND versus no intervention was: -2.67% (95% CI: -3.51, -1.85) and the corresponding RR was 0.94 (95% CI: 0.92, 0.96). Compared to NNR23, the estimated RD for the HND was -1.68% (95% CI: -2.75, -0.62) and the RR was 0.96 (95% CI: 0.93, 0.98) (**Table 3**).

**Table 3.**
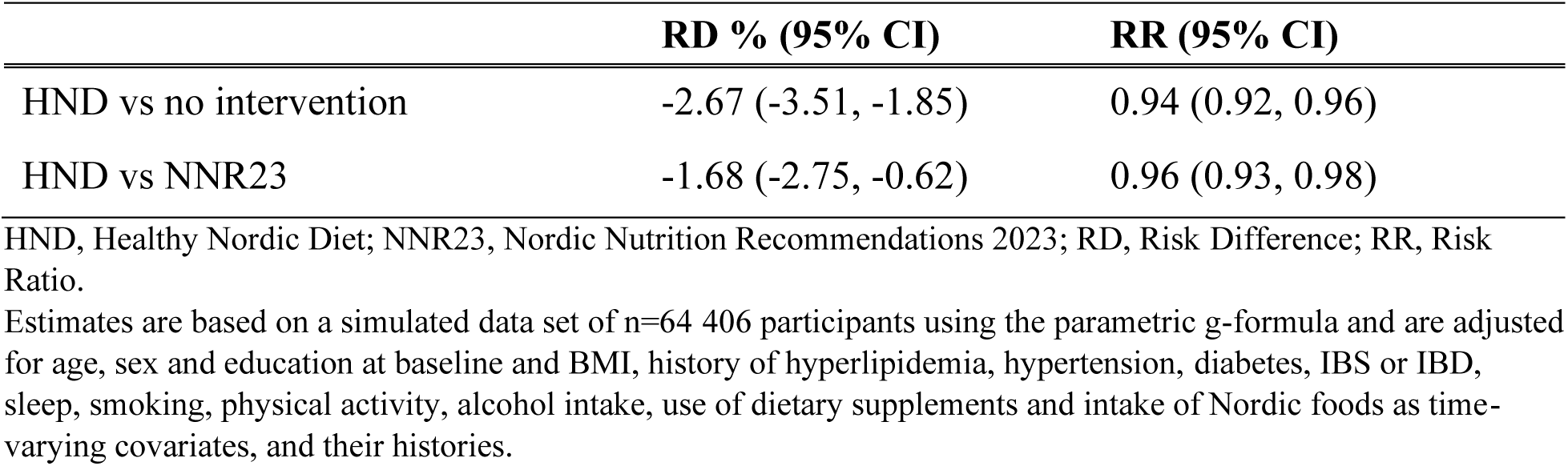
The HND and risk of all-cause mortality.

## Secondary analyses

In secondary analyses on MALO, comparing the two Nordic diets versus a low-adherence diet group, estimated RDs were -1.50% (95% CI: -9.53, -0.05) for the HND and -1.33% (95% CI: - 9.50, 0.15) for the NNR23 diet. Corresponding RRs were 0.26 (95% CI: 0.07, 0.92) and 0.34 (95% CI: 0.07, 1.26), respectively (**Table 4**). When including reducing alcohol intake to <6 grams/day as a hypothetical intervention, RDs for the HND versus no intervention and the NNR23 diet versus no intervention were -0.19% (95% CI: -0.35, -0.04) and -0.05% (95% CI: - 0.15, 0.04), respectively. Corresponding RRs were 0.70 (95% CI: 0.49, 0.93) and 0.92 (95% CI: 0.77, 1.06). Compared to NNR23, the estimated RD for the HND was -0.14% (95% CI: -0.31, 0.04) and the RR was 0.76 (95% CI: 0.53, 1.07) (**Table 5**).

**Table 4.** Nordic diets vs low-adherence diet group and risk of MALO.

|  | **RD % (95% CI)** | **RR (95% CI)** |
| --- | --- | --- |
| HND vs low-adherence group<sup>1</sup> | -1.50 (-9.53, -0.05) | 0.26 (0.07, 0.92) |
| NNR23 vs low-adherence group<sup>1</sup> | -1.33 (-9.50, 0.15) | 0.34 (0.07, 1.26) |

HND, Healthy Nordic Diet; MALO, Major Adverse Liver Outcomes; NNR23, Nordic Nutrition Recommendations 2023; RD, Risk Difference; RR, Risk Ratio.
<sup>1</sup>The low-adherence diet group was defined as high intakes of red and processed red meat, and low intakes of fruits and vegetables and whole-grain cereals. Threshold interventions for these foods were defined as reversing the signs of current dietary recommendations laid out in the NNR 2023 (e.g., if a participant consumed >500 grams of fruits and vegetables/day (in accordance with NNR 2023), the value for this individual was set to exactly 500). The estimated absolute risk in the low-adherence group was 2.03% (95% CI: 0.60, 10.23).

**Table 5.** Nordic diets with reduction in alcohol included as a threshold intervention and risk of MALO.

|  | **RD % (95% CI)** | **RR (95% CI)** |
| --- | --- | --- |
| HND with low alcohol<sup>1</sup> vs no intervention | -0.19 (-0.35, -0.04) | 0.70 (0.49, 0.93) |
| NNR23 with low alcohol<sup>1</sup> vs no intervention | -0.05 (-0.15, 0.04) | 0.92 (0.77, 1.06) |
| HND with low alcohol vs NNR23 with low alcohol | -0.14 (-0.31, 0.04) | 0.76 (0.53, 1.07) |

HND, Healthy Nordic Diet; MALO, Major Adverse Liver Outcomes; NNR23, Nordic Nutrition Recommendations 2023; RD, Risk Difference; RR, Risk Ratio.
<sup>1</sup>The population-specific threshold intervention for alcohol was set to <0.5 units (or 6 grams) of alcohol/day, corresponding to 3.5 glasses of wine/week or 14 cl of spirits/week.

Compared to no intervention, estimates were similar when intervening on all foods but one at a time, except for rapeseed oil in the HND and for whole-grain cereals and coffee in the NNR23 diet (**Figure 2**). Similar estimates as for the primary analyses were observed for the other secondary analyses (**Supplementary Table 9 and 10**).

**Figure 2.**
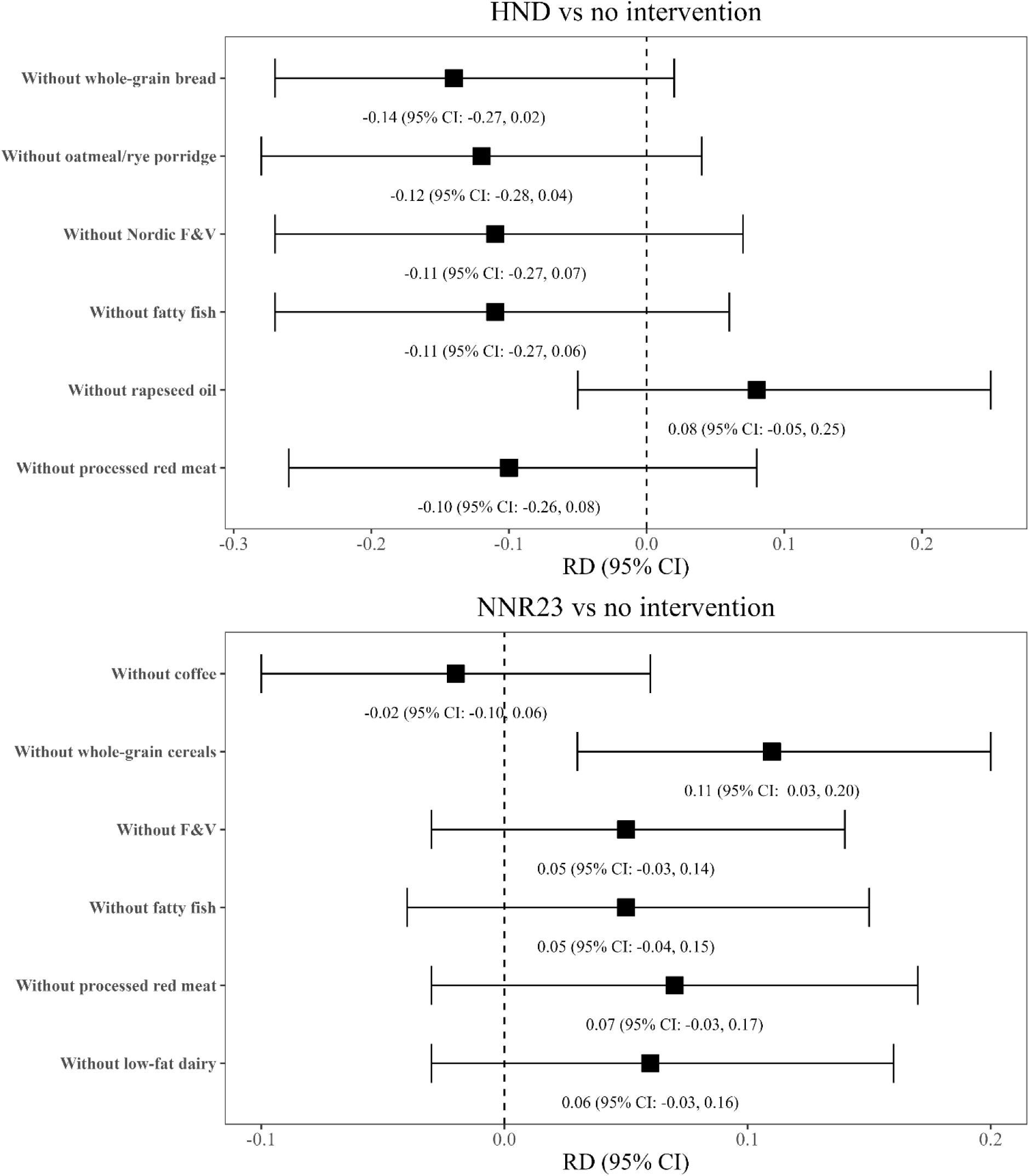
Nordic diets and MALO, excluding one food item at a time. Estimates are based on a simulated data set of n=64 406 participants using the parametric g-formula and are adjusted for age, sex and education at baseline and BMI, history of hyperlipidemia, hypertension, diabetes, IBS or IBD, sleep, smoking, physical activity, alcohol intake, use of dietary supplements and intake of Nordic foods as time-varying covariates, and their histories. HND, Healthy Nordic Diet; MALO, Major Adverse Liver Outcomes; NNR23, Nordic Nutrition Recommendations 2023; RD, Risk Difference.

Treatment effect heterogeneity was investigated for the HND using causal survival forests with baseline dietary data on the 12-year mean survival time of MALO and all-cause mortality. ATE estimates for MALO ranged from -0.007 years (95% CI: −0.021, 0.006) for effect score 1 to 0.005 years (95% CI: −0.006, 0.022) for effect score 3. ATE estimates for all-cause mortality ranged from 0.063 years (95% CI: −0.006, 0.132) for effect score 1 to 0.162 years (95% CI: 0.069, 0.256) for effect score 3 (**Supplementary Table 12**). For all-cause mortality, effect score 3 was characterized by a slightly higher proportion of women and individuals over 65 years of age with hypertension and diabetes, as well as a lower proportion of individuals with a university degree, those exercising at least two hours per week, and those with lower alcohol consumption, compared to effect score 1 and effects core 2 (**Supplementary Table 12**).

## Sensitivity analyses

Estimates from the primary analyses were similar across several sensitivity analyses examining robustness to unobserved and residual confounding, selection bias and model misspecification (**Table 6**).

**Table 6.** Sensitivity analyses for the primary analysis.

|  | HND vs no intervention |  | NNR23 vs no intervention |  | HND vs NNR23 |  |
| --- | --- | --- | --- | --- | --- | --- |
|  | RD %<br>(95% CI) | RR<br>(95% CI) | RD %<br>(95% CI) | RR<br>(95% CI) | RD %<br>(95% CI) | RR<br>(95% CI) |
| Main analysis | -0.11<br>(-0.27, 0.07) | 0.83<br>(0.59, 1.11) | 0.06<br>(-0.04, 0.15) | 1.09<br>(0.94, 1.22) | -0.17<br>(-0.38, 0.05) | 0.76<br>(0.53, 1.08) |
| Change in order<br>of covariates<sup>1</sup> | -0.11<br>(-0.27, 0.08) | 0.83<br>(0.59, 1.11) | 0.06<br>(-0.04, 0.16) | 1.09<br>(0.94, 1.23) | -0.17<br>(-0.38, 0.05) | 0.77<br>(0.53, 1.08) |
| Change in<br>functional forms<br>of covariates<sup>2</sup> | -0.11<br>(-0.27, 0.07) | 0.83<br>(0.58, 1.10) | 0.06<br>(-0.03, 0.15) | 1.09<br>(0.94, 1.22) | -0.17<br>(-0.37, 0.06) | 0.76<br>(0.53, 1.10) |
| TEI adjustment<sup>3</sup> | -0.12<br>(-0.28, 0.05) | 0.81<br>(0.57, 1.08) | 0.05<br>(-0.05, 0.14) | 1.07<br>(0.92, 1.21) | -0.17<br>(-0.37, 0.05) | 0.76<br>(0.53, 1.08) |
| MICE<sup>4</sup> | -0.11<br>(-0.28, 0.08) | 0.84<br>(0.60, 1.12) | 0.07<br>(-0.03, 0.16) | 1.10<br>(0.95, 1.23) | -0.17<br>(-0.38, 0.06) | 0.76<br>(0.53, 1.10) |
| Negative control<br>outcome<sup>5</sup> | -0.01<br>(-0.11, 0.13) | 0.97<br>(0.74, 1.35) | 0.03<br>(-0.04, 0.12) | 1.08<br>(0.88, 1.26) | -0.04<br>(-0.21, 0.15) | 0.90<br>(0.62, 1.44) |

HND, Healthy Nordic Diet; MI, Multiple Imputation; NNR23, Nordic Nutrition Recommendations 2023; RD, Risk Difference; RR, Risk Ratio; TEI, Total Energy Intake.
<sup>1</sup>Food variables and variables representing consequences of lifestyle habits (i.e., BMI, hypertension, hypercholesterolemia, diabetes and IBS/IBD) changed order.
<sup>2</sup>Variables specified as absorbing were set to binary and variables specified as zero-inflated normal were set to normal.
<sup>3</sup>Total energy intake (TEI) may work as a proxy for any unobserved confounders strongly correlated with TEI. However, adjusting for TEI imposes isocaloric comparisons.
<sup>4</sup>Multivariate imputation of chained equations (MICE) (n=1 imputation) was used to impute missing values of smoking, physical activity, sleep, use of supplements and BMI over follow-up.
<sup>5</sup>External causes of mortality excluding self-harm (e.g., transportation accidents and unintentional injuries).

## Discussion

Our target trial emulation of two population-adapted Nordic diets suggested no clear reductions in the 24-year risk of MALO in this middle aged to elderly Swedish population, when compared with each other or with no intervention. However, when compared to a low-adherence diet group and when including reducing alcohol as a hypothetical intervention, meaningful reductions in the risk of MALO from the HND were estimated. In addition, meaningful reductions in the risk of all-cause mortality were estimated for the HND when compared with both no intervention and the NNR23 diet.

Although point estimates were accompanied by wide confidence intervals, indicating no clear risk reductions, our results on the HND from our primary analysis (i.e., the direction of the estimates) combined with results from the secondary analyses support findings from a previous 12-month Swedish RCT in patients with T2D or prediabetes [6]. Compared with NNR 2012, the HND reduced liver fat, liver function markers, body weight, HbA1c, lipid markers, and C-reactive protein, several of which have been proposed as mediators on the causal pathway between diet, MALO, and all-cause mortality [39,40]. In the present study, we also estimated reductions in risk of all-cause mortality from a HND when compared with the NNR23 diet. A greater reduction in risk of MALO, although with low precision, was further estimated for the HND over the NNR23 diet when compared with a low-adherence diet group. The latter contrast, showing an estimated absolute risk reduction of 1.50%, illustrates the potential effect on MALO had individuals who currently do not meet the recommendations for F&V, red and processed red meat, and whole-grain cereals shifted their diets towards a HND. According to the most recent national dietary survey in Sweden (Riksmaten 2010–2011), only 17% of the population adhered to the recommendation for F&V, 12% for whole-grain cereals, while more than 50% consumed over 420 grams of red and processed red meat (excluding sausages) per week [41]. Our findings therefore suggest that a significant proportion of the Swedish population could benefit from adopting a HND to prevent MALO. When including reducing alcohol as a hypothetical intervention, stronger reductions in risk were estimated for the HND as compared to our main analyses, again suggesting that alcohol is an important contributor to the development of more advanced stages of liver diseases. The low precision of the estimates in our primary analysis may be due to the relatively low absolute risk of MALO, combined with the fact that more pronounced dietary shifts are required to detect clear risk reductions of MALO in an otherwise generally healthy population. This was also supported by the finding of a lower risk when comparing the HND to a low-adherence diet group.

Given the increasing incidence of several potentially lifestyle-modifiable advanced liver diseases in Sweden since the early 2000s [7], combined with the relatively poor long-term survival prognosis of MALO and the absence of long-term RCTs investigating the effects of a HND on MALO and all-cause mortality, our overall findings provide novel insights that add to the growing body of evidence supporting the HND, for population-health. Potential advantages of the HND over the NNR23 diet may partly reflect differences in emphasized food sources, such as increased use of rapeseed oil in the HND and restricted coffee intake in the NNR23 diet. When hypothetically intervened on all but one food item at a time, rapeseed oil (HND) and coffee (NNR23) emerged as key contributors to the estimated effects in the main analysis, as the point estimates changed directions when excluding these two foods from the analyses. Interestingly, both rapeseed oil and coffee have been suggested to benefit liver health in previous studies, potentially through anti-lipogenic, antioxidative, and/or anti-fibrotic mechanisms [32,33]. Whole-grain cereals also emerged as a key food component in the NNR23 diet, supporting evidence of a potentially protective effect from whole-grains on liver health [27,42].

Whether lack of clear estimated average treatment effects on MALO in our primary analysis may mask any subgroup-specific effects is a question worthy to investigate, and is most often done in the field of precision nutrition. Traditionally, subgroup-specific dietary effects, or CATEs, are estimated using a one-variable-at-a-time approach [20]. Although informative, this approach carries several methodological disadvantages compared to newer causal machine learning approaches that aim to maximize heterogeneous treatment effects by integrating multiple high-dimensional effect modifiers at once [20]. In this study, we used causal survival forests to estimate CATEs over a set of prespecified effect modifiers using baseline dietary data on the 12-year mean survival time of MALO and all-cause mortality for the HND. No clear treatment effect heterogeneity was observed for MALO. For all-cause mortality, we estimated a longer mean survival time for effect score 3 (0.162 years) compared to the other two effect scores (0.063 years and 0.073 years), suggesting that certain groups of individuals may benefit more from a HND than other groups. Interestingly, compared with effect score 1 and 2, effect score 3 was characterized by a slightly higher proportion of women and individuals over 65 years of age with hypertension and diabetes, as well as by a lower proportion of individuals with a university degree, those exercising at least two hours per week, and those with lower alcohol consumption (**Supplementary Table 12**). Whether such characteristics could be used to tailor dietary advice in practice requires replication in other cohorts, preferably including dietary variables as potential effect modifiers. The latter was not possible in our study due to lack of information on pre-baseline diet. Furthermore, future well-powered studies should aim to integrate causal machine learning methods with the target trial framework to better answer questions on whether effects from a HND may vary across specific individual characteristics.

There are several key parts that distinguish this analysis from conventional nutritional epidemiological studies, including 1) clear protocol specifications of eligibility criteria, dietary interventions and their comparators as well as causal contrasts, 2) appropriate handling of time-varying covariates in the presence of exposure-confounder feedback, using the parametric g-formula, 3) modelling, instead of treating competing events as censoring events, to estimate total effects of the diets (the censoring approach targets a direct effect and relies on even stronger assumptions), 4) estimating risk ratios and risk differences over hazard ratios and 5) performance of several sensitivity analyses to examine robustness to the underlying causal assumptions, such as the use of negative control outcomes for unobserved and residual confounding [37]. These distinctions should be regarded as key strengths of this study. For comparison, results from conventional analyses using pooled logistic regression, comparing high versus low adherence to the diets, are shown in **Supplementary Table 11**. Although the odds ratio estimates were in the same direction as the RRs estimated under the hypothetical interventions, the former estimates cannot be interpreted causally as the effects of sustained adherence to Nordic dietary patterns versus no intervention on the risk of MALO, due to reasons outlined above.

Although the target trial emulation approach aims to emulate a hypothetical pragmatic randomized trial, there are several points that need to be considered in order for our effect estimates to be interpreted causally. First, due to the inability of randomizing participants in an observational setting, unobserved and residual confounding may still be present. Our negative control outcome analysis showed, however, no clear estimated effects between diet groups and external causes of death, suggesting that we may have adjusted for all important confounders. Effect estimates were also robust when we changed the functional forms of included covariates or included total energy intake as a proxy for any unmeasured confounders strongly associated with energy homeostasis. Second, the parametric g-formula relies on strong assumptions of no model misspecifications [19]. However, changing the order of covariates or using multiple imputation to account for missing values over follow-up showed similar results as our main analysis. Further, the observed non-parametric natural course risk estimate was close to the parametric risk estimate from the g-formula, suggesting (but not guaranteeing) the absence of gross model misspecification (**Supplementary Figure 3**). Third, measurement errors of diet may have biased our estimates in any direction, depending on the structure of the errors [43,44]. However, the FFQs used in the two cohorts have been thoroughly validated, showing moderate correlations for several Nordic foods when compared with 7-day weighed food records [22,23]. Fourth, our analysis relies on strong unverifiable assumptions of sustained dietary adherence between each 10-year follow-up, which may not be realistic. Lastly, findings from this study must be interpreted within the context of the hypothetical interventions specified in this population. The thresholds for both the HND and the NNR23 diet were population-adapted to reduce the risk of positivity violations [12]. As such, we excluded nuts and vegetable oil as potential targets in the NNR23 diet, which may have contributed to the lack of estimated effects on MALO. Vegetable oils, in particular sunflower- and rapeseed oil, have been shown to decrease liver fat and improve liver function markers when compared to other plant-based oils in strictly controlled randomized trials [33,45,46]. One must also acknowledge that the no intervention group represents the population-specific distributions of each food and results may therefore be difficult to transport to other populations where these distributions differ [47].

In conclusion, we estimated no clear risk reductions from a population-adapted HND or a NNR23 diet on the 24-year risk of MALO when compared to each other or no intervention. However, when either compared to a low-adherence group or when including reducing alcohol as a hypothetical intervention or when specifying all-cause mortality as the outcome, we estimated meaningful risk reductions following the HND. Findings from these analyses therefore compliment results from randomized trials with intermediate outcomes, help to strengthen current dietary guidelines and offer new insights into the potential benefits that the HND may have on preventing MALO and all-cause mortality in the Swedish population.

## Supporting information

TARGET checklist

## Author contributions

The authors’ responsibilities were as follows – MF: initiated the idea of the project; MF: CJM, HH, AÅ, SN, DBI: designed the research; MF: analyzed the data; MF: wrote the first draft of the manuscript; MF: had primary responsibility for final content; and all authors: read and approved the final manuscript.

## Data availability

Data underlying this article cannot be shared publicly because the data are classified as sensitive data under the European General Data Protection Regulation (GDPR). Data are however available from SIMPLER (www.simpler4health.se) for researchers who meet the criteria, that is, an ethical approval is demanded, for access to SIMPLER data.

## Acknowledgements

We would like to thank the National Research Infrastructure SIMPLER (www.simpler4health.se), supported by the Swedish Research Council (2017-00644 and 2021-00160), for providing us with databases from the Swedish Mammography Cohort (SMC) and the Cohort of Swedish Men (COSM). A special thanks to Anna-Karin Kolseth for her support in this project. We would also like to acknowledge the National Academic Infrastructure for Supercomputing in Sweden (NAISS) at Uppsala Multidisciplinary Center for Advanced Computational Science (UPPMAX), partially funded by the Swedish Research Council (2022-06725).

## Funding

MF and this work was supported by a research grant from the Danish Diabetes and Endocrine Academy, which is funded by the Novo Nordisk Foundation, grant number NNF22SA0079901. DBI was funded by the Steno Diabetes Center Aarhus, Aarhus University Hospital, which is partially funded by an unrestricted donation from the Novo Nordisk Foundation. HH was supported by grants from The Swedish Research Council, The Swedish Cancer Society, Region Stockholm (CIMED and clinical researcher award), The Swedish Heart and Lung Foundation and others.

## Conflicts of interests

HH:s institutions have received research funding from Astra Zeneca, EchoSens, Gilead, Intercept, MSD, Novo Nordisk, Takeda and Pfizer with HH as the PI. He has served as consultant, speaker or on advisory boards for Astra Zeneca, Boehringer Ingelheim, Bristol Myers-Squibb, GSK, Echosens, Ipsen, MSD and Novo Nordisk and has been part of hepatic events adjudication committees for Arrowhead, Boehringer Ingelheim, KOWA and Jazz Pharma. No other authors declare any conflicts of interests.

## Notes

### Author Declarations

SMC and COSM were conducted in accordance with the declaration of Helsinki and were approved by the Regional Ethical Review Board at Karolinska Institutet, Stockholm, Sweden.

