## Supplementary Material for "Estimating the Effects of Nordic Diets on the Risk of Major Adverse Liver Outcomes: a Target Trial Emulation across Two Cohorts in Sweden"

[1]Department of Clinical Medicine, Aarhus University, Aarhus, Denmark.
[2]Steno Diabetes Center Aarhus, Aarhus University Hospital, Aarhus, Denmark.
[3]Department of Public Health and Caring Sciences, Uppsala University, Uppsala, Sweden.
[4]Novo Nordisk A/S.
[5]Institute of Environmental Medicine, Karolinska Institutet, Stockholm, Sweden.
[6]Department of Medicine, Huddinge, Karolinska Institutet, Stockholm, Sweden.
[7]Division of Hepatology, Department of Upper GI Diseases, Karolinska University Hospital, Stockholm, Sweden.
[8]Department of Public Health, Aarhus University, Aarhus, Denmark.

**Table of contents**

Threshold interventions for the Nordic diets……….……………………………………..2

Categorization of foods……………………………………………………………3-4

Diagnostic codes for liver diseases……………………………..……………………5-6

Specification of covariates…….……………………………………………………..7

Habitual diet across time points………………………………………………………….8

Baseline characteristics stratified by adherence…………………………………………9

Proportion intervened on……………………..……………………………………….10

Secondary analyses……………………………………...…………………………….11

Conventional analyses…………………………………………………………………...12

Causal survival forest……………………………………………………………...13-15

Study flow-chart……………………………………………..……………...……16

DAG for the main analysis………………………………………………………………17

Parametric vs non-parametric covariate estimates……………………………………….18

Excerpt of R-code for main analysis…………………………………..………..19-21

**Supplementary Table 1.** Threshold interventions for the two Nordic diets.

| | HND | | NNR23 | |
|---|---|---|---|---|
| **Foods** | **Original thresholds** | **Adapted thresholds** | **Original thresholds** | **Adapted thresholds** |
| Coffee | - | - | ≤ 4 cups/day | Same |
| Whole-grain cereals | - | - | ≥ 90 grams/day | Same |
| F&V | - | - | ≥ 500 grams/day | ≥ 250 grams/day |
| Pulses | - | - | ≥ 100 grams/day | ≥ 10 grams/day |
| Fatty fish | ≥ 3 portions/week | ≥ 0.5 portions/week | ≥ 2 portions/week | ≥ 0.5 portions/week |
| Other fish | - | - | ≥ 1 portion/week | ≥ 0.5 portions/week |
| Red meat | ≤ 280 grams/week | ≤ 420 grams/week | ≤ 280 grams/week | ≤ 420 grams/week |
| Processed red meat | ≤ 70 grams/week | ≤ 350 grams/week | ≤ 70 grams/week | ≤ 350 grams/week |
| Low-fat dairy | - | - | ≥ 350 grams/day | ≥ 100 grams/day |
| Potatoes | - | - | ≥ 130 grams/day | Same |
| Egg | - | - | ≤ 1 egg/day | Same |
| Poultry | - | - | ≤ 350 grams/week | Same |
| Juice | - | - | ≤ 100 grams/day | Same |
| Nuts | - | - | ≥ 20 grams/day | Excluded* |
| Vegetable oil | - | - | ≥ 25 grams/day | Excluded* |
| Whole-grain bread | ≥ 2 portions/day | Same | - | - |
| Oatmeal/rye porridge | ≥ 3 portions/week | ≥ 0.5 portions/week | - | - |
| Nordic F&V | ≥ 250 grams/day | ≥ 125 grams/day | - | - |
| Rapeseed oil | Yes in cooking and salad dressing | Yes in cooking or salad dressing | - | - |

F&V, Fruits and vegetables; HND, Healthy Nordic Diet; NNR23, Nordic Nutrition Recommendations 2023.
*Few participants consumed >0 grams of nuts and vegetable oil, hence the exclusion of these foods from the hypothetical interventions.

**Supplementary Table 2.** Categorizations of foods across the three time points.

| Foods | 1997 | 2009 | 2019 |
|---|---|---|---|
| Coffee | Coffee (unspecified) | Filtered/instant coffee + Unfiltered/other coffee | Filtered/instant coffee + Unfiltered/other coffee |
| Whole-grain cereals | Whole grain/meal bread + Crisp bread + Oatmeal porridge + Wheat or oat bran | Wholegrain pasta/spaghetti/macaroni + Wholegrain rice + Fiber enriched bread + Whole grain/meal bread + Crisp bread + Oatmeal/rye porridge + Wheat or oat bran | Wholegrain pasta/spaghetti/macaroni + Wholegrain rice + Fiber enriched bread + Whole grain/meal bread + Crisp bread + Oatmeal/rye porridge + Wheat or oat bran |
| F&V | Spinach + Lettuce/iceberg lettuce + Cabbage (white, red, Chinese) + Cauliflower + Broccoli/brussels sprouts + Carrots + Beetroot + Garlic + Onion/leek + Tomatoes/tomato juice + Bell pepper + Mixed vegetables + Orange/citrus fruits + Apple/pear + Banana + Other fruits + Berries (fresh or frozen) | Spinach + Lettuce/iceberg lettuce + Cabbage (white, red, Chinese) + Cauliflower + Broccoli/brussels sprouts + Carrots + Beetroot + Garlic + Onion + Leek + Tomatoes/tomato juice + Bell pepper + Avocado + Olives + Corn + Mixed frozen vegetables + Other vegetables + Orange/citrus fruits + Apple/pear + Banana + Other fruits + Berries (fresh or frozen) | Spinach + Lettuce/iceberg lettuce + Cabbage (white, red, Chinese) + Cauliflower + Broccoli/brussels sprouts + Carrots + Beetroot + Garlic + Onion + Leek + Tomatoes/tomato juice + Bell pepper + Avocado + Olives + Corn + Mixed frozen vegetables + Other vegetables + Orange/citrus fruits + Apple/pear + Banana + Other fruits + Berries (fresh or frozen) |
| Pulses | Pea soup/brown beans/beans/lentils | Pea soup + Beans/lentils/chick peas | Pea soup + Beans/lentils/chick peas |
| Fatty fish | Salmon/whitefish/red char + Herring/mackerel | Salmon + Herring/mackerel + Tuna + Sardines | Salmon + Herring/mackerel + Tuna |
| Other fish | Cod/saithe/fish fingers | Smoked fish + Cod/saithe/plaice/grenadier + Pike/perch + Other fish | Smoked fish + Cod/saithe/plaice/grenadier + Pike/perch + Other fish |
| Red meat | Minced meat dishes + Pork (steak/casserole) + Beef/veal (steak/casserole) + Liver/kidney | Minced meat dishes + Pork (steak/casserole) + Beef/veal (steak/casserole) + Other meat + Liver/kidney | Minced meat dishes + Pork (steak/casserole) + Beef/veal (steak/casserole) + Other meat + Liver/kidney |
| Processed red meat | Sausage (unspecified) + Blood pudding/sausage + Liver pâté (unspecified) + Cold cuts, meat/sausage | Bacon + Sausage + Lean sausage + Other sausage + Blood pudding/sausage + Liver pâté (not low-fat) + Low-fat liver pâté + Cold cuts, meat (e.g., ham/turkey) + Cold cuts, sausage (e.g. salami) | Bacon + Sausage + Lean sausage + Other sausage + Blood pudding/sausage + Liver pâté (not low-fat) + Low-fat liver pâté + Cold cuts, meat (e.g., ham/turkey) + Cold cuts, sausage (e.g. salami) |
| Low-fat dairy | Semi-skimmed milk (1.5% fat) + Skimmed milk (0.5% fat) + Low-fat sour milk/yoghurt (≤0.5% fat) + Low-fat crème fraiche + Low-fat cheese | Semi-skimmed milk (1.5% fat) + Skimmed milk (0.5% fat) + Reduced fat sour milk (1.5% fat) + Low-fat sour milk/yoghurt (≤0.5% fat) + Low-fat crème fraiche + Low-fat cream/sour cream + Low- | Semi-skimmed milk (1.5% fat) + Skimmed milk (0.5% fat) + Reduced fat sour milk (1.5% fat) + Low-fat sour milk/yoghurt (≤0.5% fat) + Low-fat crème fraiche + Low-fat |

| | | | |
|---|---|---|---|
| | | fat hard cheese + Low-fat cheese spread | cream/sour cream + Low-fat hard cheese + Low-fat cheese spread |
| Potatoes | Boiled potatoes + Fried potatoes | Boiled potatoes + Fried potatoes + Baked/mashed potatoes | Boiled potatoes + Fried potatoes + Baked/mashed potatoes |
| Vegetable oil | Soft/table margarine + Low-fat margarine + Cooking oil | Soft/table margarine + Low-fat margarine + Margarine + Oil, as bread spread | Soft/table margarine + Low-fat margarine + Margarine + Oil, as bread spread |
| Egg | Eggs/omelet | Eggs/omelet | Eggs/omelet |
| Poultry | Chicken/other poultry | Chicken/other poultry | Chicken/other poultry |
| Juice | Orange/grapefruit juice | Orange/grapefruit juice | Orange/grapefruit juice |
| Whole-grain bread | Whole grain/meal bread + Crisp bread | Fiber enriched bread + Whole grain/meal bread + Crisp bread | Fiber enriched bread + Whole grain/meal bread + Crisp bread |
| Oatmeal/rye porridge | Oatmeal porridge | Oatmeal/rye porridge | Oatmeal/rye porridge |
| Nordic F&V | Apple/pear + Berries (fresh or frozen) + Cabbage (white, red, Chinese) + Cauliflower + Broccoli/brussels sprouts + Carrots + Beetroot | Apple/pear + Berries (fresh or frozen) + Cabbage (white, red, Chinese) + Cauliflower + Broccoli/brussels sprouts + Carrots + Beetroot | Apple/pear + Berries (fresh or frozen) + Cabbage (white, red, Chinese) + Cauliflower + Broccoli/brussels sprouts + Carrots + Beetroot |
| Nuts | Nuts/almonds (unspecified) | Nuts/almonds + Peanuts | Nuts/almonds + Peanuts |
| Rapeseed oil | Rapeseed oil in cooking and rapeseed oil in home-made dressing | Rapeseed oil in cooking and rapeseed oil in home-made dressing | Rapeseed oil in cooking and rapeseed oil in home-made dressing |

F&V, Fruits and vegetables.

**Supplementary Table 3.** Diagnostic codes for the outcome and other chronic liver diseases.

| Diagnosis | ICD-8 | ICD-9 | ICD-10 |
|---|---|---|---|
| **MASLD[1]** | | | |
| MASLD | NA | 571.8 | K76.0 |
| MASH | NA | NA | K75.8 |
| **Other chronic liver diseases (CLD)[1]** | | | |
| ALD | 571,00, 571,01 | 571.0-571.3 | K70 |
| Chronic viral hepatitis | 999,2, 070 | 070 | B18-B19 |
| Autoimmune liver disease | NA | 571.6, 576.1 | K83.0A, K74.3, K75.4 |
| Hemochromatosis | 273,2 | 275.0 | E83.1 |
| Wilson | 273,3 | 275.1 | E83.0B |
| Alpha-1 antitrypsin deficiency | NA | 277.6 | E88.0A, E88.0B |
| Budd-Chiari | NA | 453.0 | I82.0, K76.5 |
| Chronic hepatitis, unspecified | 570 | 571.4 | K73.9, K73.2 |
| Secondary or unspecified biliary cirrhosis | NA | 571.6 | K74.4, K74.5 |
| Unspecified cirrhosis | 571,9 | 571.5 | K74.6 |
| Gastric varices, not bleeding | NA | NA | I86.4 |
| Esophageal varices, not bleeding | NA | 456.1, 456.21 | I85.9, I98.2 |
| **Decompensated cirrhosis (MALO outcome)[1]** | | | |
| Portal hypertension | 571,9 | 572.3 | K76.6 |
| Ascites[2] | 785.3 | 789.5 | R18 |

| | | | |
|---|---|---|---|
| Hepatorenal syndrome | NA | 572.4 | K76.7 |
| Esophageal varices, bleeding | 456.0 | 456.0, 456.20 | I85.0, I98.3 |
| **Hepatocellular carcinoma (MALO outcome)[1]** | | | |
| HCC | - | 155.0 | C22.0 (ICD-O/2 in the Swedish Cancer Register) |
| **Liver transplantation (MALO outcome)[1]** | | | |
| Transplantation status | NA | V42.7 | Z94.4 |

[1]Codes used to exclude participants with CLD at baseline.
[2]Requires a concomitant diagnostic code from any CLD except for MASLD/MASH.

**Supplementary Table 4.** Specification of time-fixed and time-varying covariates.

| Variable name | As dependent | As independent |
|---|---|---|
| **Time-fixed** | | |
| Age | Not predicted | Continuous |
| Sex | Not predicted | 2 categories |
| Education | Not predicted | 3 categories |
| **Time-varying** | | |
| **Non-dietary variables** | | |
| BMI | Linear | Continuous |
| Hypertension | Logistic, absorbing[1] | 2 categories |
| Hypercholesterolemia | Logistic, absorbing[1] | 2 categories |
| Diabetes | Logistic, absorbing[1] | 2 categories |
| IBS or IBD | Logistic, absorbing[1] | 2 categories |
| Sleep | Linear | Continuous |
| Smoking | Multinomial | 3 categories |
| Physical activity | Multinomial | 4 categories |
| Alcohol intake | Linear | Continuous |
| Use of supplements | Logistic | 2 categories |
| **Dietary variables** | | |
| Whole-grain bread | Linear | Spline (3 knots) |
| Oatmeal/rye porridge | Linear, zero-inflated normal[2] | Spline (3 knots) |
| Nordic F&V | Linear | Continuous |
| Fatty fish | Linear, zero-inflated normal[2] | Continuous |
| Rapeseed oil | Logistic | 2 categories |
| Red meat | Linear | Continuous |
| Processed red meat | Linear | Continuous |
| Coffee | Linear | Continuous |
| Whole-grain cereals | Linear | Spline (3 knots) |
| F&V | Linear | Continuous |
| Pulses | Linear, zero-inflated normal[2] | Spline (3 knots) |
| Other fish | Linear, zero-inflated normal[2] | Continuous |
| Low-fat dairy | Linear, zero-inflated normal[2] | Continuous |
| Potatoes | Linear | Continuous |
| Egg | Linear | Continuous |
| Poultry | Linear, zero-inflated normal[2] | Continuous |
| Juice | Linear, zero-inflated normal[2] | Continuous |

BMI, Body Mass Index; F&V, Fruits and vegetables; IBD, Inflammatory Bowel Disease; IBS, Irritable Bowel Syndrome.

[1]Absorbing means that a value is not allowed to further change once it has changed from 0 to 1.

[2] Zero-inflated normal first runs a logistic regression model for intake vs no intake and subsequently runs a linear regression model. The indicator for intake vs not is then multiplied by the coefficient from the linear regression model. These models are specified for variables with many zeros.

**Supplementary Table 5.** Habitual diet across time points[1].

| Food component | 1997 (n=64 406) | 2009 (n=38 809) | 2019 (n=20 748) |
|---|---|---|---|
| Coffee (g/day) | 552 (354-820) | 424 (328-656) | 368 (328-552) |
| Whole-grain bread (g/day) | 94 (53-154) | 104 (66-160) | 89 (50-137) |
| F&V (g/day) | 290 (192-422) | 317 (209-458) | 303 (197-441) |
| Pulses (g/day) | 19 (16-24) | 21 (16-33) | 21 (9-32) |
| Fatty fish (g/day) | 9 (6-15) | 18 (12-29) | 14 (8-24) |
| Other fish (g/day) | 10 (8-25) | 22 (13-33) | 21 (8-33) |
| Red meat (g/day) | 45 (28-66) | 45 (29-68) | 38 (23-59) |
| Red processed meat (g/day) | 32 (19-45) | 45 (28-68) | 39 (22-59) |
| Low-fat dairy (g/day) | 192 (19-397) | 220 (60-413) | 141 (14-305) |
| Potatoes (g/day) | 99 (65-129) | 107 (75-147) | 103 (69-135) |
| Egg (g/day) | 8 (3-16) | 10 (5-16) | 15 (5-36) |
| Poultry (g/day) | 8 (8-10) | 10 (8-29) | 9 (8-29) |
| Juice (g/day) | 14 (0-45) | 10 (0-44) | 0 (0-30) |
| Oatmeal (g/day) | 10 (0-43) | 11 (0-74) | 12 (0-95) |
| Nordic F&V (g/day) | 123 (74-185) | 122 (74-186) | 120 (68-188) |
| Whole-grain cereals (g/day) | 123 (73-218) | 165 (100-255) | 142 (80-238) |
| Rapeseed oil (n (%))[2] | 11 975 (18.6) | 20 354 (52.4) | 12 076 (58.2) |

F&V, Fruits and Vegetables.

[1]Dietary variables are presented as median (IQR) for continuous variables or counts (%) for categorical variables.

[2]Yes in cooking or in salad dressing.

**Supplementary Table 6.** Baseline characteristics stratified by dietary adherence scores[1].

| | **HND 0-4 points[2] (n=40 010)** | **HND 5-7 points[2] (n=24 396)** | **NNR23 0-10 points[2] (n=40 262)** | **NNR23 11-13 points[2] (n=24 144)** |
|---|---|---|---|---|
| Sex (n (%) women) | 15 994 (40.0) | 13 235 (54.3) | 14 814 (36.8) | 14 415 (59.7) |
| Age (years) | 58 (52-67) | 61 (54-69) | 58 (52-67) | 61 (55-69) |
| University degree (n (%) | 7083 (17.7) | 4819 (19.8) | 7153 (17.8) | 4749 (19.7) |
| Smoking (n (%) current) | 10 579 (26.4) | 4391 (18.0) | 10 808 (26.8) | 4162 (17.2) |
| Physical activity (≥2 hours/week of exercise) | 21 455 (53.6) | 15 757 (64.6) | 21917 (54.4) | 15 295 (63.4) |
| Sleep (hours/day) | 7 (7-8) | 7 (7-8) | 7 (7-8) | 7 (7-8) |
| Use of supplements (n (%) yes regularly) | 7090 (17.7) | 5615 (23.0) | 7145 (17.7) | 5560 (23.0) |
| BMI (kg/m$^2$) | 25.1 (23.1-27.5) | 24.9 (22.9-27.2) | 25.1 (23.1-27.4) | 24.9 (22.9-27.3) |
| Hypertension (n (%)) | 8493 (21.2) | 5506 (22.6) | 8291 (20.6) | 5708 (23.6) |
| Hypercholesterolemia (n (%)) | 4554 (11.4) | 3046 (12.5) | 4581 (11.4) | 3019 (12.5) |
| Type 2 diabetes (n (%)) | 1831 (4.6) | 1325 (5.4) | 1846 (4.6) | 1310 (5.4) |
| IBS or IBD (n (%)) | 231 (0.6) | 100 (0.4) | 224 (0.6) | 107 (0.4) |
| Total energy intake (kcal/day) | 2132 (1602-2785) | 2119 (1709-2672) | 2230 (1676-2891) | 1994 (1619-2498) |
| Alcohol intake (g/day) | 6 (2-12) | 5 (1-10) | 6 (2-12) | 4 (1-9) |
| Coffee (g/day) | 556 (354-848) | 531 (354-759) | 636 (384-885) | 424 (328-591) |
| Whole-grain bread (g/day) | 81 (44-147) | 106 (74-160) | 88 (50-155) | 100 (66-148) |
| F&V (g/day) | 242 (163-352) | 374 (274-502) | 242 (165-376) | 352 (269-474) |
| Pulses (g/day) | 19 (16-24) | 19 (16-51) | 19 (16-24) | 19 (16-51) |
| Fatty fish (g/day) | 8 (4-14) | 14 (8-19) | 8 (4-15) | 12 (8-18) |
| Other fish (g/day) | 10 (8-25) | 10 (8-25) | 10 (8-25) | 10 (8-25) |
| Red meat (g/day) | 50 (29-80) | 38 (25-52) | 52 (29-80) | 36 (25-50) |
| Red processed meat (g/day) | 34 (20-52) | 28 (17-39) | 34 (20-52) | 28 (18-39) |
| Low-fat dairy (g/day) | 156 (0-378) | 217 (62-408) | 134 (0-348) | 256 (136-428) |
| Potatoes (g/day) | 99 (65-135) | 98 (65-127) | 102 (73-151) | 89 (65-109) |
| Egg (g/day) | 8 (5-17) | 8 (3-16) | 8 (5-17) | 8 (3-15) |
| Poultry (g/day) | 8 (8-10) | 9 (8-10) | 8 (8-10) | 9 (8-10) |
| Juice (g/day) | 14 (0-45) | 14 (0-50) | 14 (0-50) | 11 (0-36) |
| Oatmeal/rye porridge (g/day) | 0 (0-13) | 32 (10-111) | 0 (0-33) | 13 (0-88) |
| Nordic F&V (g/day) | 96 (60-145) | 168 (130-230) | 103 (62-164) | 153 (106-213) |
| Whole-grain cereals (g/day) | 105 (54-189) | 165 (106-255) | 113 (62-215) | 144 (98-225) |
| Rapeseed oil (n (%) yes in cooking or salad dressing) | 3142 (7.9) | 8833 (36.2) | 6111 (15.2) | 5864 (24.3) |

[1]Values are presented as median (IQR) for continuous variables and as count (%) for categorical variables. [2]The HND and the NNR23 scores are based on the same thresholds as for the main emulated target trial. BMI, Body Mass Index; HND, Healthy Nordic Diet; IBD, Inflammatory Bowel Disease; IBS, Irritable Bowel Syndrome; NNR23, Nordic Nutrition Recommendations 2023.

**Supplementary Table 7.** Percent intervened on for the HND and the NNR23 diet.

| | Interpretation | HND (n=64 406) | NNR23 (n=64 406) |
|---|---|---|---|
| Baseline % intervened on | Proportion of individuals who would have been required to change their diet at baseline to adhere to the diet strategy | 97.62 | 96.32 |
| Average % intervened on | Average proportion of individuals who would have been required to change their diet at each time point to adhere to the diet strategy | 93.89 | 96.10 |
| Cumulative % intervened on | Cumulative proportion of individuals who would have been required to change their diet at any time point over follow-up to adhere to the diet strategy | 99.92 | 99.97 |

HND, Healthy Nordic Diet; NNR23, Nordic Nutrition Recommendations 2023.

**Supplementary Table 8.** Components adhered to at baseline for the HND and the NNR23 diet.

| Component | Population-adapted threshold | HND |
|---|---|---|
| Whole-grain bread | ≥ 2 portions/day | 72.8 % |
| Fatty fish | ≥ 0.5 portions/week | 70.0 % |
| Red meat | ≤ 420 grams/week | 68.9 % |
| Red processed meat | ≤ 350 grams/week | 79.9 % |
| Oatmeal/rye porridge | ≥ 0.5 portions/week | 48.7 % |
| Nordic fruits and vegetables | ≥ 125 grams/day | 49.0 % |
| Rapeseed oil | Yes in cooking or salad dressing | 18.6 % |
| All components | All of the above | 2.4 % |
| **Component** | **Population-adapted threshold** | **NNR23** |
| Coffee | ≤ 4 cups/day | 56.3 % |
| Whole-grain cereals | ≥ 90 grams/day | 66.5 % |
| Fruits and vegetables | ≥ 250 grams/day | 60.2 % |
| Pulses | ≥ 10 grams/day | 82.2 % |
| Fatty fish | ≥ 0.5 portions/week | 70.0 % |
| Other fish | ≥ 0.5 portions/week | 85.6 % |
| Red meat | ≤ 420 grams/week | 68.9 % |
| Red processed meat | ≤ 350 grams/week | 79.9 % |
| Low-fat dairy | ≥ 100 grams/day | 64.2 % |
| Potatoes | ≤ 130 grams/day | 75.3 % |
| Egg | ≤ 1 egg/day | 98.2 % |
| Poultry | ≤ 350 grams/week | 98.9 % |
| Juice | ≤ 100 grams/day | 84.2 % |
| All components | All of the above | 3.7 % |

HND, Healthy Nordic Diet; NNR23, Nordic Nutrition Recommendations 2023.

**Supplementary Table 9.** Nordic diets and risk of MALO exempting individuals who report excluding dairy or gluten from being intervened on those specific foods.

| | RD % (95% CI) | RR (95% CI) |
|---|---|---|
| HND (with exemption)[1] vs no intervention | -0.11 (-0.27, 0.08) | 0.84 (0.59, 1.12) |
| NNR23 (with exemption)[1] vs no intervention | 0.06 (-0.03, 0.15) | 1.09 (0.95, 1.22) |
| HND (with exemption)[1] vs NNR23 (with exemption)[1] | -0.16 (-0.37, 0.06) | 0.76 (0.53, 1.10) |

HND, Healthy Nordic Diet; MALO, Major Adverse Liver Outcomes; NNR23, Nordic Nutrition Recommendations 2023; RD, Risk Difference; RR, Risk Ratio.

Estimates are based on a simulated data set of n=64 406 participants using the parametric g-formula and are adjusted for age, sex and education at baseline and BMI, history of hyperlipidemia, hypertension, diabetes, IBS or IBD, sleep, smoking, physical activity, alcohol intake, use of dietary supplements and intake of Nordic foods as time-varying covariates, and their histories.

[1]Participants who reported excluding dairy or gluten from their diet were exempt from being hypothetically intervened on whole-grain bread and oatmeal/rye porridge in the HND and whole-grain cereal and low-fat dairy in the NNR23 diet over follow-up.

**Supplementary Table 10.** Nordic diets and risk of MALO changing the thresholds for low-fat dairy, potatoes and whole-grain cereals.

| | RD % (95% CI) | RR (95% CI) |
|---|---|---|
| NNR23 (with changed thresholds)[1] vs no intervention | 0.01 (-0.09, 0.10) | 1.01 (0.86, 1.15) |
| HND vs NNR23 (with changed thresholds)[1] | -0.11 (-0.33, 0.12) | 0.82 (0.55, 1.22) |

HND, Healthy Nordic Diet; MALO, Major Adverse Liver Outcomes; NNR23, Nordic Nutrition Recommendations 2023; RD, Risk Difference; RR, Risk Ratio.

Estimates are based on a simulated data set of n=64 406 participants using the parametric g-formula and are adjusted for age, sex and education at baseline and BMI, history of hyperlipidemia, hypertension, diabetes, IBS or IBD, sleep, smoking, physical activity, alcohol intake, use of dietary supplements and intake of Nordic foods as time-varying covariates, and their histories.

[1]Thresholds were changed for low-fat dairy (from ≥100 to ≤500 grams per day), potatoes (from ≤130 to ≥50 grams per day) and whole-grain cereals (from ≥90 to ≥110 grams per day) in the NNR23 diet.

### Conventional analyses

To compare our estimates from the target trial emulation against the wider literature, we conducted conventional nutritional epidemiological analyses of the two Nordic dietary patterns on the risk of MALO using pooled logistic regression. First, adherence scores for the HND and the NNR23 guidelines were constructed based on median intakes of each food (e.g., 1 point was given if a participant consumed > median intake of whole-grain bread or <median intake of red meat). The median categorization approach is a common exposure classification strategy in nutritional epidemiology. Scores from each food were summed and further divided into 0-2, 3-4 and 5-7 points for the HND and 0-5, 6-7 and 8-13 points for the NNR23 dietary pattern. A pooled logistic regression model with time as factor was specified, adjusting for the same time-fixed and time-varying confounders as in the main analysis. Robust standard errors were derived from the sandwich estimator. A last observation carried forward approach was implemented for missing values on confounders during follow-up. The results from this analysis are shown below.

**Supplementary Table 11.** Pooled logistic regression analyses of the two Nordic diets on MALO.

| | | **HND** | |
|---|---|---|---|
| | 0-2 points (ref) | 3-4 points | 5-7 points |
| Odds ratios (95% CI) | 1.00 | 0.93 (0.73-1.19) | 0.96 (0.69-1.33) |
| | | **NNR23** | |
| | 0-5 points (ref) | 6-7 points | 8-13 points |
| Odds ratios (95% CI) | 1.00 | 1.02 (0.79-1.32) | 1.08 (0.80-1.47) |

CI, Confidence Interval; HND, Healthy Nordic Diet; NNR23, Nordic Nutrition Recommendations 2023.

Odds ratios derived from the pooled logistic regression (approximating discrete time hazard ratios) differed to some degree from the risk ratios derived from the main analysis contrasting the HND and the NNR23 guidelines vs each other or no intervention. Discrepancies may be due to several reasons, including 1) different exposure contrasts, 2) competing events treated as censoring events in the conventional approach, 3) non-collapsibility of odds ratios, 4) collider bias induced by adjusting for time-varying confounders affected by prior exposure using conventional statistical approaches, and 5) adjustment for previous diet not included. As such, the above quantities cannot be interpreted as causal effects from adhering to a HND or the NNR23 guidelines vs no intervention over follow-up.

**Causal survival forest**

The ultimate goal of causal inference is to be able to estimate individual causal treatment effects**;** that is, the effect of a treatment on an outcome for each individual in the study. However, due to the so-called fundamental problem of causal inference, that we can only observe the outcome under either the treatment or control condition, but never both for the same individual, true individual treatment effects remain inherently unobservable. This essential missing data problem has led to numerous attempts to approximate individual treatment effects through subgroup analyses based on a set of characteristics, including genetic, demographic, anthropometric, and more recently, multi-omics variables.

While stratified subgroup analyses can provide valuable insights, they are subject to important limitations. Statistically, conducting multiple subgroup analyses increases the risk of both type I and type II errors due to multiple comparisons and reduced power. More importantly perhaps, from a patient perspective, heterogeneous treatment effects (HTE) likely arise from interactions across multiple covariates simultaneously, rather than from a single characteristic in isolation. For example, if one dietary study reports a beneficial treatment effect on liver function in individuals with type 2 diabetes (T2D) but a harmful effect in males, what dietary advice would a patient presented as male with T2D receive? This ambiguity highlights the limitations of the one-variable-at-a-time approach to HTE.

A promising solution to this challenge is the application of machine learning (ML) methods in causal inference, such as causal forests, which allow for estimation of conditional average treatment effects (CATEs) based on high-dimensional baseline covariate information (i.e., $\tau(x) = E[Y_i^{(1)} – Y_i^{(0)} \mid X_i=x]$, whereby $Y^{(1)}$ is the potential outcome under treatment and $Y^{(0)}$ is the potential outcome under no treatment. X is a vector of covariates). These methods can estimate CATEs while flexibly modeling complex interactions and nonlinearities. Estimated CATEs can subsequently be summarized, for instance, by stratifying individuals into quantiles of predicted treatment effects, often referred to as effect scores, to examine patterns of effect heterogeneity across these scores. In this secondary analysis, we first grouped individuals into a high (5-7 points) or a low (0-4 points) adherence group of the HND based on the intervention thresholds from the main analysis. We then restricted the sample to baseline intake only and followed participants until event, death or administrative end of follow-up, whichever occurred first and calculated time in study by years. This analysis was performed for both MALO and all-cause mortality for the HND only.

The full study sample was then randomly divided into a training set (50%) and an estimation set (50%). CATEs for the 12-year mean survival time (RMST) were estimated non-parametrically using honest causal survival forests, thereby reducing overfitting and enabling valid inference under the assumption of exchangeability (i.e., that the diet groups are comparable at baseline or from counterfactual theory; that the potential outcomes are independent of the treatment). The forest was trained using 5000 trees with honest splitting, a 50 % sample fraction, horizon=12 and default tuning parameters in the *grf*-package in R. Potential effect modifiers included were all identified confounders in the main analysis of the target trial emulation.

In short, a causal forest builds trees based on splits of the covariates that maximize treatment effect heterogeneity, leaving both treated and controls in separate so-called leaves. These trees, and thus leaves, are then aggregated into a forest and averaged to estimate how treatment effects vary across individuals. The average treatment effect (ATE) of the HND on MALO and all-cause mortality, i.e., $ATE = E[Y^{(1)} - Y^{(0)}]$, was estimated using the doubly-robust Augmented Inverse Propensity Weighting (AIPW) estimator. Subsequently, ATEs in tertiles (from now on called effect scores (ES)) of ranked CATEs were calculated using the AIPW estimator and the distribution of each covariate was examined descriptively between each ES.

**Supplementary Table 12.** Average treatment effects and descriptive data on potential effect modifiers across effect scores[1].

| | HND and MALO (5-7 points vs 0-4 points) | | |
|---|---|---|---|
| | **ES1** | **ES2** | **ES3** |
| RMST ATE (years (95% CI))[1] | -0.007 (−0.021, 0.006) | 0.009 (-0.001, 0.019) | 0.005 (-0.009, 0.019) |
| Age (years) | 59 (16) | 59 (14) | 59 (15) |
| Age>65 years (%) | 38.9 | 27.5 | 36.6 |
| Sex (% women) | 46.3 | 44.0 | 45.9 |
| Education (% University) | 17.4 | 19.9 | 18.2 |
| BMI (kg/m$^2$) | 25.0 (4.4) | 25.1 (4.4) | 25.1 (4.3) |
| Obesity (%) | 9.9 | 10.4 | 9.9 |
| Hypertension (%) | 22.6 | 20.9 | 21.7 |
| Hypercholesterolemia (%) | 11.6 | 12.1 | 11.7 |
| Diabetes (%) | 5.2 | 4.8 | 4.7 |
| IBS or IBD (%) | 0.5 | 0.5 | 0.5 |
| PA (% ≥2 hours/week of exercise) | 57.6 | 57.3 | 58.4 |
| Use of supplements (% regularly) | 20.4 | 18.9 | 20.0 |
| Smoking (% current) | 22.4 | 24.0 | 23.2 |
| Sleep (hour/days) | 7 (1) | 7 (1) | 7 (1) |
| Alcohol intake (g/day) | 4.9 (9.5) | 5.5 (9.8) | 5.2 (9.6) |
| >1 unit/day of alcohol (%) | 21.5 | 23.6 | 22.2 |
| | **HND and all-cause mortality (5-7 points vs 0-4 points)** | | |
| | **ES1** | **ES2** | **ES3** |
| RMST ATE (years (95% CI))[1] | 0.063 (−0.006, 0.132) | 0.073 (0.009, 0.136) | 0.162 (0.069, 0.256) |
| Age (years) | 59 (15) | 59 (14) | 59 (17) |
| Age>65 years (%) | 33.7 | 26.7 | 42.3 |
| Sex (% women) | 45.3 | 43.5 | 47.3 |
| Education (% University) | 18.6 | 20.1 | 16.7 |
| BMI (kg/m$^2$) | 25.1 (4.4) | 25.0 (4.4) | 25.0 (4.5) |
| Obesity (%) | 10.4 | 9.9 | 9.9 |
| Hypertension (%) | 21.3 | 20.2 | 23.7 |
| Hypercholesterolemia (%) | 11.5 | 12.3 | 11.6 |
| Diabetes (%) | 4.8 | 4.5 | 5.4 |
| IBS or IBD (%) | 0.5 | 0.5 | 0.5 |
| PA (% ≥2 hours/week of exercise) | 58.4 | 57.1 | 53.5 |
| Use of supplements (% regularly) | 19.4 | 18.9 | 20.8 |
| Smoking (% current) | 23.2 | 24.1 | 22.4 |
| Sleep (hour/days) | 7 (1) | 7 (1) | 7 (1) |
| Alcohol intake (g/day) | 5.3 (9.7) | 5.6 (9.9) | 4.7 (9.2) |
| >1 unit/day of alcohol (%) | 22.7 | 23.9 | 20.7 |

Descriptive values are presented as median (IQR) for continuous variables and as % for categorical variables.
[1]Restricted mean survival times (RMST) are estimated over 12 years of follow-up using causal survival forests with n=5000 trees and baseline dietary data. Estimates are presented in years. Positive signs indicate longer time free from MALO or longer time of survival whereas negative signs indicate shorter time free from MALO or shorter time of survival. The ATE for MALO in the full estimation sample was 0.002 years (95% CI: −0.005, 0.010) comparing high versus low adherence to the HND whereas the ATE for all-cause mortality was 0.099 years (95% CI: 0.055, 0.143). ATE, Average Treatment Effect; BMI, Body Mass Index; ES, Effect Score; HND, Healthy Nordic Diet; IBD, Inflammatory Bowel Disease; IBS, Irritable Bowel Syndrome; MALO, Major Adverse Liver Outcomes; PA, Physical Activity; RMST, Restricted Mean Survival Time.

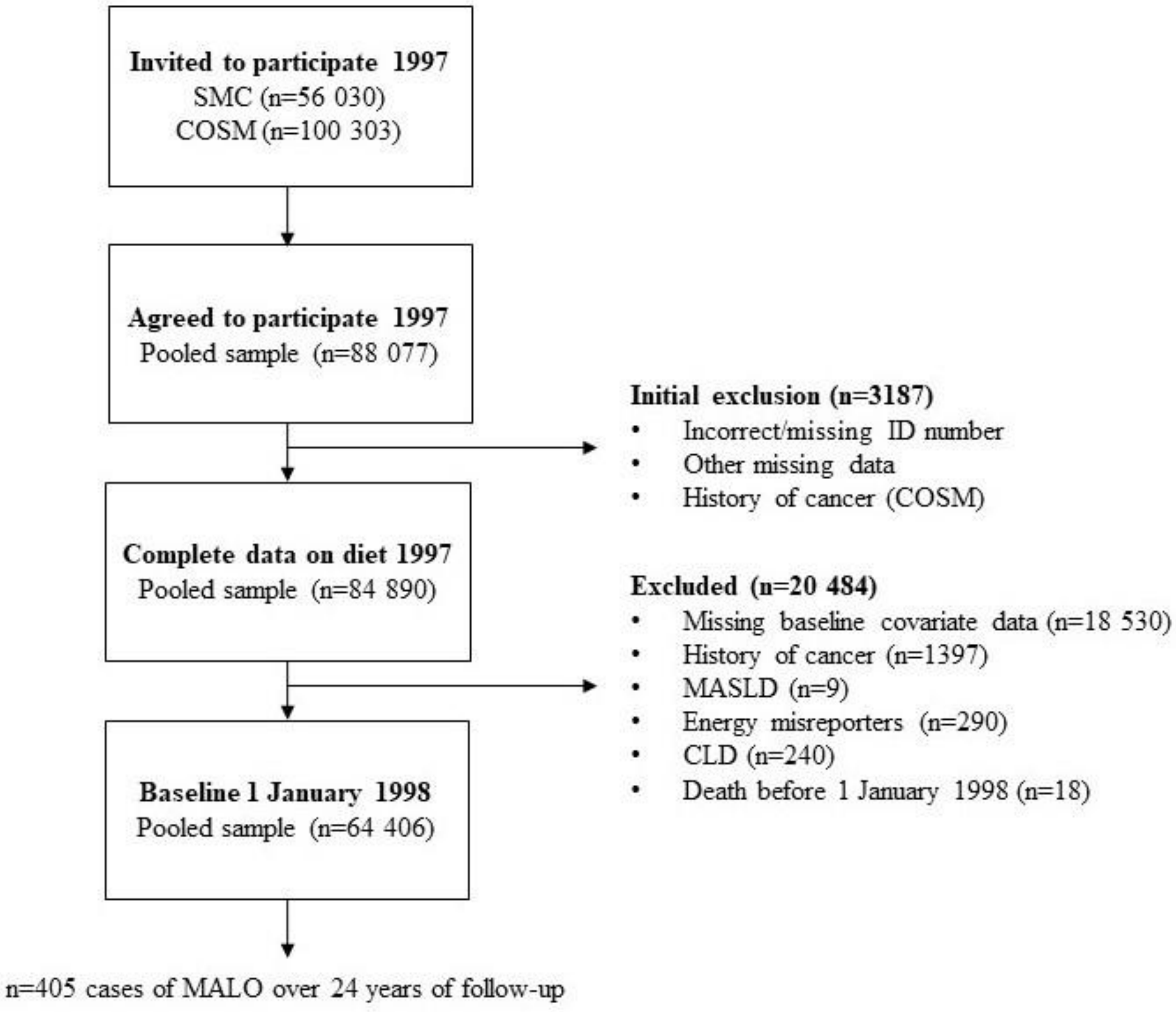


**Supplementary Figure 1.** Study flow-chart of the pooled cohorts.
Missing data on baseline covariates were: n=9362 for physical activity, n=6818 for use of supplements, n=3258 for BMI, n=1529 for smoking, n=1523 for sleep and n=539 for education. CLD, Chronic Liver Disease; COSM, Cohort of Swedish Men; MALO, Major Adverse Liver Outcomes; MASLD, Metabolic-dysfunction Associated Steatotic Liver Disease; SMC, Swedish Mammography Cohort.

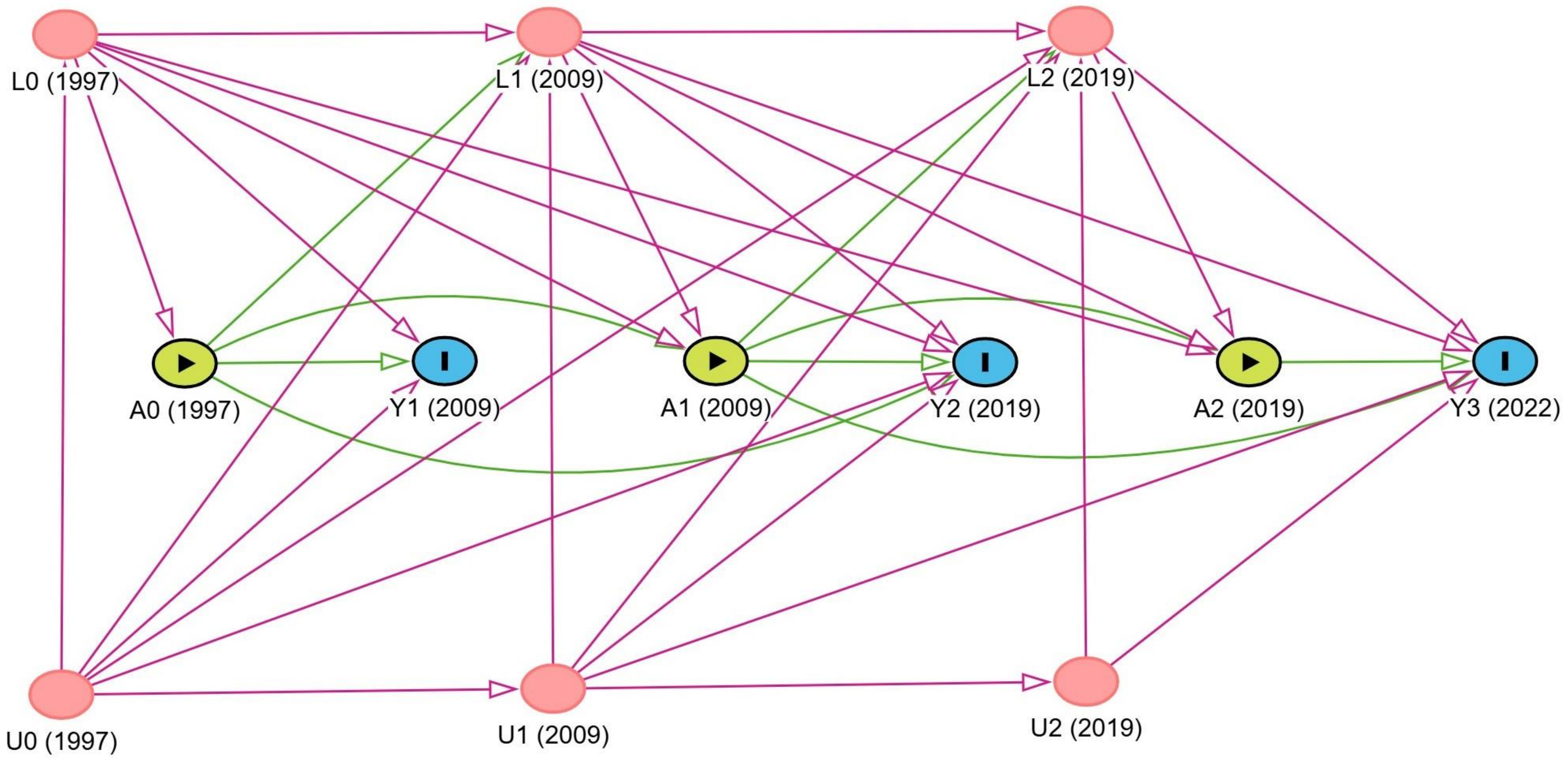


**Supplementary Figure 2**. A simplified directed acyclic graph (DAG) depicting our causal assumptions of included variables. A0-A2 represent the exposure, Y1-Y3 the outcome, L0-L2 both time-fixed and time-varying confounders (e.g., education as a time-fixed confounder and BMI as a time-varying confounder) and U0-U2 unobserved time-fixed and time-varying confounders over the three time points. Adjusting for time-varying confounding in the presence of exposure-confounder feedback (e.g., A0 (1997) → L1 (2009) → A1 (2009)) requires the use of g-methods, such as the parametric g-formula. Using conventional methods such as outcome regression adjustment would partly block the pathway from A0 (1997) → L1 (2009) → Y2 (2019) but also induce collider bias through A0 (1997) → L1 (2009) ← U1 (2009) → Y2 (2019).

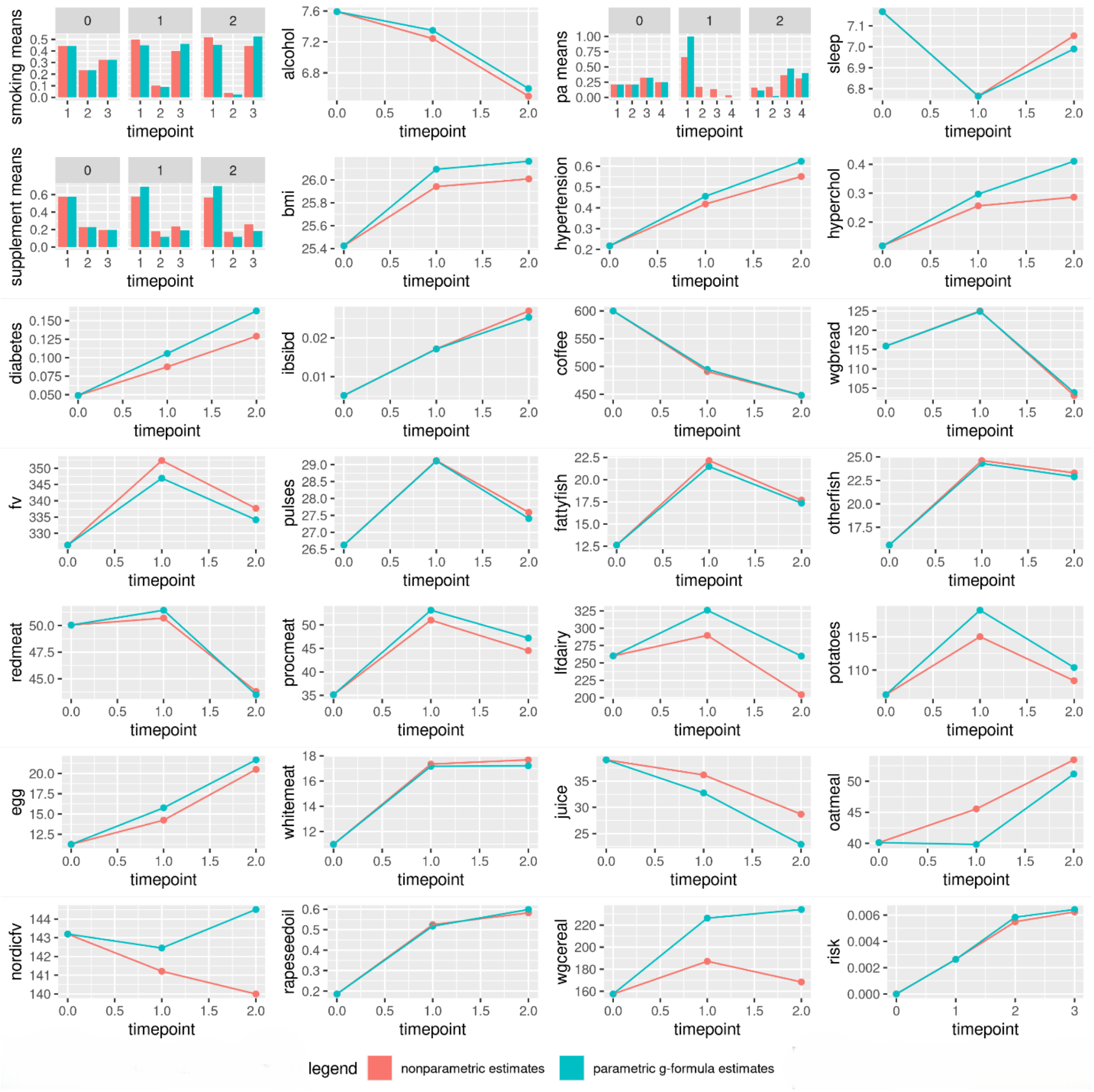


**Supplementary Figure 3.** Parametric g-formula vs non-parametric natural course covariate mean estimates for the main analysis. The non-parametric risk estimate (0.0062) is close to the parametric g-formula risk estimate (0.0064), supporting (but not guaranteeing) the absence of gross model misspecification.

**R-code**

```
#Main purpose of original script: overall setup and running the parametric g-formula
#The script is an excerpt of the original script
#The script only includes 3 threshold interventions for each diet, 3 time-fixed and 2 time-varying confounders
#The script is for educational purposes
#Author: Michael Fridén

#—————————————————————————————————————————————

#Load packages
library(dplyr)
library(gfoRmula)
library(Hmisc)

#—————————————————————————————————————————————

#Load data in long format
malo_long < -readRDS("/castor/project/home/micfri/nordic_diets_malo/data/malo_long.Rds")

#—————————————————————————————————————————————

#LOCF for NA over follow-up
vars_to_fill <- c("bmi")

malo_long <- malo_long %>%
arrange(SIMPKEY, timepoint) %>%
group_by(SIMPKEY) %>%
fill(all_of(vars_to_fill), .direction = "down") %>%
ungroup()

#—————————————————————————————————————————————

#Analysis
#Overall setup
id <- "SIMPKEY"
time_points <- 3 #1997, 2009 and 2019
time_name <- "timepoint"
nsimul <- 64406 #Number of simulations = number of participants in the pooled cohort
ncores <- as.numeric(parallel::detectCores())

#Intervention variables
intvars <- list( c("wgbread", "redmeat", "nordicfv"), #HND
c("wgcereal", "redmeat", "fv") #NNR23
)
```

```
#Threshold interventions
interventions <- list( list( #Intervention 1: HND
c(threshold, 60, Inf), #wgbread >= 60 grams/day (2 servings/day)
c(threshold, -Inf, 60), #redmeat <= 60 grams/day (420 grams/week)
c(threshold, 125, Inf) #nordicfv >= 125 grams/day
),
list( #Intervention 2: NNR23
c(threshold, 90, Inf), #wgcereal >= 90 grams/day
c(threshold, -Inf, 60), #redmeat <= 60 grams/day (420 grams/week)
c(threshold, 250, Inf) #fv >= 250 grams/day
) )

#Description of the interventions
int_descript <- c("HND", "NNR23")

#Outcome (MALO)
outcome_type <- "survival"
outcome_name <- "event"
ymodel <- event ~ age + sex + education + bmi + lag1_bmi + hypertension + lag1_hypertension
+ Hmisc::rcspline.eval(wgbread, nk=3) + lag1_wgbread + redmeat + lag1_redmeat + nordicfv +
lag1_nordicfv + Hmisc::rcspline.eval(wgcereal, nk=3) + lag1_wgcereal + fv + lag1_fv + timepoint

#Competing event (death from other causes)
compevent_name <- "death"
cmodel <- death ~ age + sex + education + bmi + lag1_bmi + hypertension + lag1_hypertension
+ Hmisc::rcspline.eval(wgbread, nk=3) + lag1_wgbread + redmeat + lag1_redmeat + nordicfv +
lag1_nordicfv + Hmisc::rcspline.eval(wgcereal, nk=3) + lag1_wgcereal + fv + lag1_fv + timepoint

#Covariates
#Covtype absorbing is specified for variables that, once taken 1, never takes 0 again
#Covtype zero-inflated normal is specified for variables with many zeros
basecovs <- c("age", "sex", "education")
covnames <- c("bmi", "hypertension", "wgbread", "redmeat", "nordicfv", "wgcereal", "fv")
covtypes <- c("normal", "absorbing", "zero-inflated normal", "normal", "normal", "zero-inflated
normal", "normal")
histories <- c(lagged)
histvars <- list(c("bmi", "hypertension", "wgbread", "redmeat", "nordicfv", "wgcereal", "fv"))

#Covariate models
covparams <- list(covmodels = c(
bmi ~ age + sex + education + lag1_bmi + lag1_hypertension + lag1_wgbread + lag1_redmeat +
lag1_nordicfv + lag1_wgcereal + lag1_fv + timepoint,

hypertension ~ age + sex + education + bmi + lag1_bmi + lag1_wgbread + lag1_redmeat +
lag1_nordicfv + lag1_wgcereal + lag1_fv + timepoint,
```

```
wgbread ~ age + sex + education + bmi + lag1_bmi + hypertension + lag1_hypertension +
lag1_wgbread + lag1_redmeat + lag1_nordicfv + lag1_wgcereal + lag1_fv + timepoint,

redmeat ~ age + sex + education + bmi + lag1_bmi + hypertension + lag1_hypertension +
Hmisc::rcspline.eval(wgbread, nk=3) + lag1_wgbread + lag1_redmeat + lag1_nordicfv +
lag1_wgcereal + lag1_fv + timepoint,

nordicfv ~ age + sex + education + bmi + lag1_bmi + hypertension + lag1_hypertension +
Hmisc::rcspline.eval(wgbread, nk=3) + lag1_wgbread + redmeat + lag1_redmeat + lag1_nordicfv
+ lag1_wgcereal + lag1_fv + timepoint,

wgcereal ~ age + sex + education + bmi + lag1_bmi + hypertension + lag1_hypertension +
Hmisc::rcspline.eval(wgbread, nk=3) + lag1_wgbread + redmeat + lag1_redmeat + nordicfv +
lag1_nordicfv + lag1_wgcereal + lag1_fv + timepoint,

fv ~ age + sex + education + bmi + lag1_bmi + hypertension + lag1_hypertension +
Hmisc::rcspline.eval(wgbread, nk=3) + lag1_wgbread + redmeat + lag1_redmeat + nordicfv +
lag1_nordicfv + Hmisc::rcspline.eval(wgcereal, nk=3) + lag1_wgcereal + lag1_fv + timepoint)
)
#————————————————————————————————————————

#Running the parametric g-formula with competing events (n=500 bootstrap samples)
#No intervention arm is the reference group
gform_main_comp <- gformula(obs_data = malo_long, id = id, outcome_name =
outcome_name, outcome_type = outcome_type, ymodel = ymodel, compevent_name =
compevent_name, compevent_model = cmodel, time_points = time_points, time_name =
time_name, covnames = covnames, covparams = covparams, covtypes = covtypes, basecovs =
basecovs, intvars = intvars, interventions = interventions, int_descript = int_descript, histories =
histories, histvars = histvars, ref_int = 0, #change ref_int to 2 for NNR23 as reference group
nsimul = nsimul, parallel = TRUE, nsamples = 500, ncores = ncores, seed = 1234)

gform_main_comp

#————————————————————————————————————————
```
